# Risk factors for suicide attempts in community alcohol treatment

**DOI:** 10.1101/2024.04.29.24306528

**Authors:** John Robins, Katherine Morley, Richard Hayes, Amelia Jewell, Nicola Kalk

## Abstract

**Background:** Alcohol Use Disorder is associated with suicide and suicide attempts, and addiction treatment services have a role in suicide prevention. We aimed to identify risk factors for suicide attempt among a cohort of community-based alcohol treatment service users.

**Methods:** Linked data from 4415 adults accessing secondary addiction services for alcohol treatment between 2006 and 2019 in London, UK, were used to identify risk factors for suicide attempt. Cox proportional hazards regression estimated the relative increase or decrease in hazard associated with each risk factor on a composite outcome event; death by suicide or contact with emergency psychiatric care within one year of starting treatment.

**Findings:** There were 468 (10.5%) crisis care contact events, and <10 suicide deaths. After adjustment, factors associated with increased hazard of crisis care contact or death by suicide were history of suicide attempt (HR 1.83[1.43-2.33]), poor mental health (HR 1.81[1.41-2.32]), current suicidal ideation (HR 1.65[1.18-2.31]), use of drugs other than cocaine, cannabis and opiates (HR 1.41[1.02-1.95]), female sex (HR 1.34[1.10-1.65]) and social isolation (HR 1.24[1.02 - 1.51]). Factors associated with reduced hazard of crisis care contact or death by suicide were alcohol abstinence (HR 0.51[0.31-0.83], ref>30 units), drinking 1-15 units (HR 0.64[0.49-0.85], ref>30 units), Black ethnicity (HR 0.61[0.45-0.83]) and living with children (HR 0.74[0.56-0.99]).

**Interpretation:** The identified risk factors for suicide attempt can help risk formulation and safety planning among patients accessing alcohol treatment services.

**Funding:** National Institute for Health Research Biomedical Research Centre at South London and Maudsley NHS Foundation Trust, King’s College London.

**Research in context:** *Evidence before this study:* Alcohol Use Disorder (AUD) is a risk factor for suicide and suicide attempt, and among people with AUD those accessing addiction treatment services are particularly at risk. Effective suicide risk formulation and safety planning requires an understanding of the demographic, clinical and circumstantial factors that are associated with increased or decreased risk of suicide attempt in the population of people accessing treatment for their alcohol use. We searched PubMed using search terms ((("alcohol use disorder") OR ("alcohol depend*") OR ("substance use disorder") AND (("treat*") OR ("service*"))) AND (("suicide attempt") OR ("suicid*"))) as well as Google Scholar and cited reference searching in Web of Science, to identify previous studies of risk factors for suicidal behaviour in cohorts engaged with some form of Substance Use Disorder (SUD) treatment which included alcohol at least to a minimal degree, and which measured a suicide-related outcome after treatment commencement. The impact of the risk factors identified in these studies varied greatly, reflecting heterogeneity in the substance use profiles and settings of the samples used. We found no longitudinal studies which consider risk factors for suicidal behaviour in a purely alcohol-using sample accessing community-based addiction care. The single consistent risk factor for suicidal behaviour across these studies was a previous history of suicide attempt.

*Added value of this study:* Our study uses 14 years’ worth of structured data from service users accessing Community Drug and Alcohol Team (CDAT) treatment primarily for their alcohol use. A range of risk factors for suicide attempt (measured via contact with crisis care services) or death by suicide in the year following treatment start were identified: predisposing factors included a history of suicide attempt, female sex and White ethnicity; modifiable factors included social isolation, poor mental health, current suicidal ideation or carer concern, and use of drugs other than cocaine, cannabis and opiates; protective factors included abstinence from or relatively low use of alcohol, and children living with the service user. This is the first prospective analysis of risk factors for suicidal behaviour in a purely alcohol-using sample accessing community-based addiction care. This population represent the largest proportion of CDAT service use, with a uniquely elevated suicide risk.

*Implications of all the available evidence:* A wide range of risk factors for suicide and suicide attempt can be identified among people accessing alcohol treatment, providing population-specific contextual knowledge that can aid patient-centred suicide assessment and safety planning, and a potential framework within which potential avenues for intervention can be identified.

## Introduction

Alcohol use is a risk factor for suicide and suicide attempt (1,2). The risk of death by suicide among people with Alcohol Use Disorder (AUD) is 9 times higher in men and 16 times higher in women compared to the general population (3). Among people with AUD, estimates of suicidal behaviour are particularly elevated in treatment-seeking samples; a meta-analysis of 31 global studies of patients treated for AUD reported a crude suicide mortality rate of 2.36/1000 person-years, accounting for 7.34% of all deaths (4). Another compared eleven studies that reported rates of suicide attempt among adults who had used drug or alcohol treatment services and estimated a sample size weighted mean of 24.9% reporting a lifetime suicide attempt (5). This is approximately ten times higher than the 2.7% estimate of lifetime suicide attempt in the general population from WHO surveys (6), and approximately 3 times higher than the wider population with AUD (7). A recent analysis of all suicide deaths between October 2021 and September 2022 in England and Wales found that 8% (n=428) were by people who had been in contact with drug and alcohol services within the year prior to their death, almost half of whom had been seeking treatment primarily for alcohol (8).

Given such prevalence of suicidal behaviour among those treated for AUD, it is vital that drug and alcohol treatment services are equipped to formulate and manage suicide risk. Suicide risk assessment and management strategies have moved away from attempting to predict and stratify suicide risk in individuals, towards understanding the suicide risk factors that are pertinent to particular populations, which can be used to inform the collaborative process of personalised safety planning with individual service users (9,10). The profile of suicide risk factors in people seeking treatment for AUD may be different to those of the wider population. For example, criminal justice system involvement is associated with a two to three times increased risk for suicide in general population samples (11), but among a sample of US military veterans engaging with substance use treatment, it was found to be a protective factor against suicide (12). The protective effect of parenthood on suicide risk is well-established in general population samples (13) but few data exist around whether having children is a protective factor among those seeking treatment for AUD.

Previous observational studies of risk factors for suicidal behaviour in treatment-seeking AUD patients have been conducted, but have been limited by: small sample sizes (14); a lack of prospective design with outcomes restricted to either recent pre-treatment suicidal behaviour (15,16) or lifetime suicidal behaviour (17,18); the role of alcohol dependence either not being considered (19) or comprising a low percentage of overall cohort (20,21); and sample generalisability being limited by cohorts comprised of subgroups of the treatment seeking population, e.g. those accessing inpatient or residential rehabilitation and thus representing only the very severe end of the AUD spectrum (22,23). There are no longitudinal studies which consider risk factors for suicidal behaviour in a purely alcohol-using sample accessing community-based addiction care.

**Supplementary Table 1** contains a summary of the risk factors for suicidal behaviour identified by previous longitudinal studies involving cohorts engaged with some form of Substance Use Disorder (SUD) treatment which included alcohol at least to a minimal degree, and which measured a suicide-related outcome after treatment commencement. The findings from these studies have been largely inconsistent. The only reliably identified risk factor is a previous suicide attempt, with the associated increase in odds of later suicide attempt ranging from three-fold (23) to eight-fold (22), after adjustment. Suicidal intent at baseline was found to increase risk of suicide attempt within three years in a cohort of primarily heroin users (OR 2.24, 1.09-4.60) (24), yet no effect was found after adjustment in the only other study in which it was reported (25). The effect of sociodemographic traits and social circumstances is also ambiguous; after adjustment Pavarin et al. (2021) found being separated or divorced to be the only social risk factor in their study associated with later death by suicide (IRR 2.13, 1.05-4.33) (26), and Darke et al. (2007) found a strong association between social isolation and suicide (OR 4.26, 2.21-10.27) (24). However, other studies found no effect associated with living without a partner (27), marital status (23), or social support (28). The effect of non-alcohol drug use is similarly inconsistent; three studies found no effect of cocaine use on later suicide (8,26) or suicide attempt (22) whereas two studies estimating odds of suicide attempt within a year found a three-fold increase compared to other drug use (28) and a 2% increase for every year of lifetime cocaine use (12). Most recently, a case-control study by NCISH compared characteristics of people who died by suicide within 12 months of contact with a drug and alcohol service with matched living controls, and found that people who died by suicide were more likely to have used alcohol (OR 2.77, 2.22-3.45) and less likely to have used heroin (OR 0.33, 0.25-0.42) (8).

### Current study

The current study aims to identify different clinical, demographic, and social risk factors for suicide attempt—defined here as either fatal (i.e. death by suicide) or non-fatal (i.e. contact with emergency psychiatric care)—among a cohort of secondary alcohol treatment service users in London, UK. Although all treatment-seeking alcohol users present a higher risk of suicidal crisis than the general population, it is hypothesised that that risk of suicidal behaviour is not uniform in this cohort.

Understanding the profile of suicide risk factors specific to this population will help clinicians to collaborate with service users in effective and preventative safety planning. This study will be the first longitudinal study of risk factors for suicidal behaviour in individuals seeking treatment for their alcohol use in community-based secondary addiction treatment in the UK.

## Methods

### Design and setting

This prospective cohort study uses routinely recorded electronic health record data from a cohort of patients accessing Community Drug and Alcohol Team (CDAT) alcohol treatment. CDAT services provide specialist community care for those with substance addictions; in England, alcohol is the most common problem substance among those starting CDAT treatment, with 64% reporting a problem with alcohol, two thirds of whom report no other problem substance (29). The CDAT services used in this study were from the four London boroughs where the South London and Maudsley NHS Trust (SLaM) provides secondary addictions and mental health services.

### Data sources

The data used were drawn from information recorded by CDAT staff on the National Drug Treatment Monitoring System (NDTMS). The NDTMS is a series of administrative datasets which provide structured data on all engagements with secondary addictions treatment services in England. NDTMS data from SLaM-run CDAT services are available to researchers via the SLaM Biomedical Research Centre Clinical Record Interactive Search (CRIS) application. The design, operation and development of CRIS has been described elsewhere (30). Mortality data were obtained via linkage to Office for National Statistics (ONS) cause-of-death data, which is taken from an individual’s death certificate.

The ONS regularly provides cause-of-death data to the SLaM Clinical Data Linkage Service which acts as a ‘trusted third party’ to case-match pseudonymised CRIS data with the ONS data via NHS number. This linked data is then stripped of the original CRIS identifier and returned to the researcher for analysis.

### Ethical approval

Ethical approval for the study was granted via the Oxford C Research Ethics Committee, reference (18/SC/0372), which covers all uses of CRIS as an anonymised database for secondary analysis.

Specific approval from the CRIS oversight committee was granted under Project 20-030 ‘The impact of alcohol treatment on suicidal crisis and use of emergency psychiatric care’. All patient identifiable information was removed prior to use by the CRIS application. All data remained within the NHS firewall during analysis. Frequencies fewer than 10 are suppressed in this manuscript as per CRIS guidelines.

### Participants

All adult treatment episodes from 1^st^ January 2006 accepted on to CDAT structured treatment were eligible for inclusion. Episodes were excluded if they related to other branches of addiction treatment, such as inpatient treatment, outreach, or stop-smoking services. Episodes which did not entail any engagement with the service were excluded, i.e., episodes which were closed on the same day as they were opened, with a recorded discharge reason of ‘non-attendance’. The latest date of inclusion was 28th Feb 2019, to allow for at least a full year of follow-up for all participants before the impact of COVID-19 on service engagement and mortality (31).

Further exclusions were made after examining the data, including the removal of ‘impossible episodes’ where the discharge date preceded the start date, and ‘inherited episodes’ where data had been transferred in bulk onto CRIS after local service recommissioning and no accurate episode start dates were available.

Only treatment episodes which were related to alcohol use were included, identified by alcohol listed as a ‘problem substance’ in NDTMS fields. Opioid users and cases involving injecting drug use were removed due to their high potential for confounding, due to the greatly increased risk of death from any of multiple potential causes found in these groups (32,33) and the difficulty in disambiguating intent in cases of opioid overdose (34). Service users who use opioids and alcohol are treated under the opioid pathway in CDAT services (29).

Only episodes which had a recorded risk assessment—indicating presence or absence of a suicide attempt history and/or current suicidal ideation—were included. Repeat episodes of the same individual were also excluded to avoid bias by repeat measurement. The final dataset contained 4451 records (**Figure 1**).

**Figure 1.**
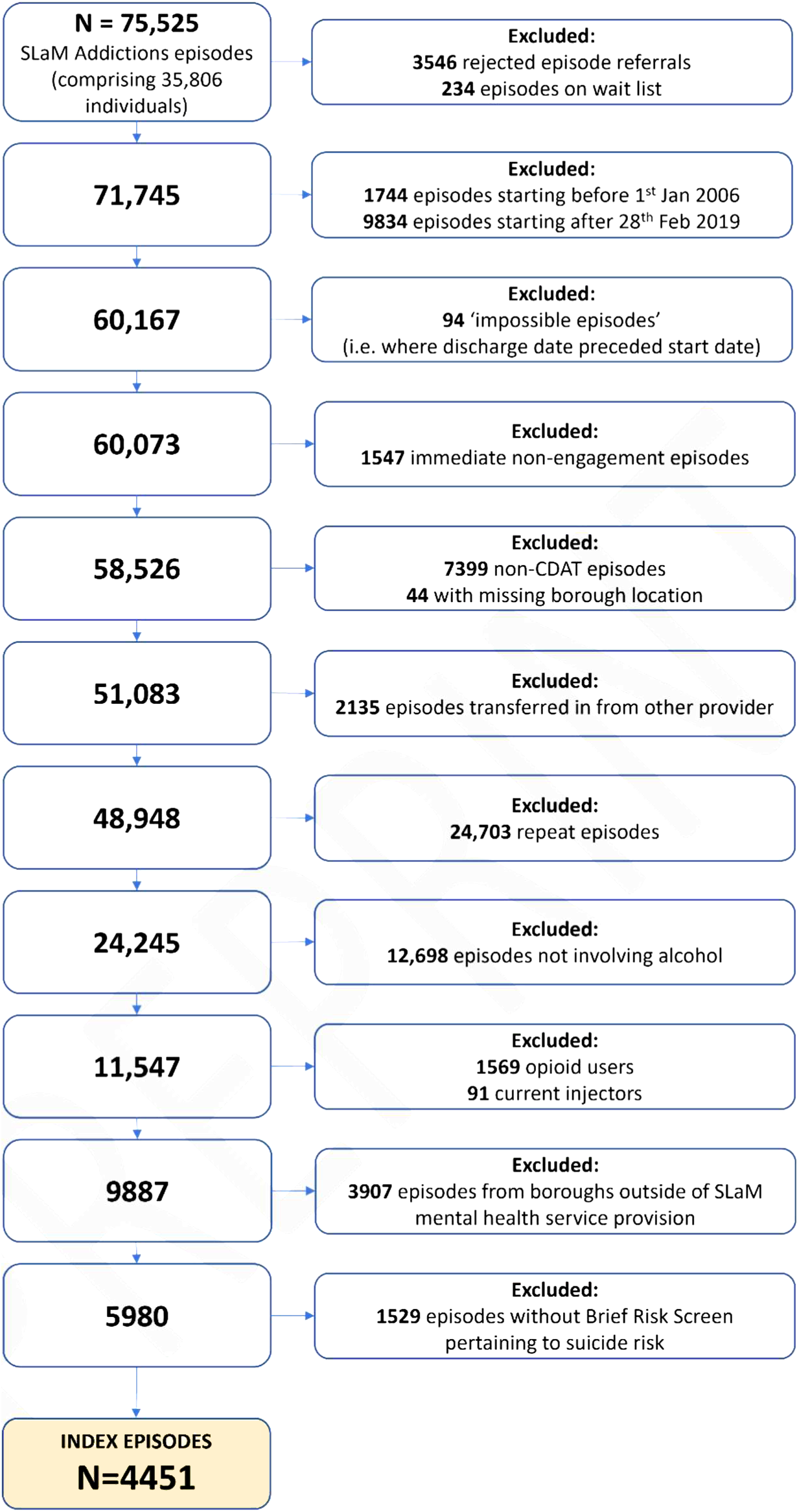
Application of exclusion criteria to extracted data

### Measures

Apart from cause-of-death data, all measures were derived from structured fields on CRIS, including reference data recorded at treatment start (35), Treatment Outcome Profile (TOP) data, and Addictions Brief Risk Screen (BRS-A) data. The TOP is a well-validated instrument which includes patient-reported outcomes from the previous 28 days, recorded at treatment start and at least every 6 months throughout treatment (36). The BRS-A is a structured risk assessment tool used by addiction services in SLaM that is standardly recorded at treatment start. OHID guidance requires that the first TOP is completed +/- 14 days from the first contact. As such, all records from within 14 days either side of index date were included; where more than one record existed for a particular service user, the record closest to the index date was used. The index date was defined as the date the treatment episode was accepted on to the CDAT caseload. **Supplementary Table 2** contains further details on the derivation of each measure.

### Exposure: Risk factors

#### Suicide risk

Individuals were classed as having a history of suicide attempt if a ‘yes’ was recorded for the BRS-A item *History of suicide attempts*. Individuals were classed as having current suicidal ideation if there was a ‘yes’ recorded for either or both BRS-A items pertaining to suicidal intent at the point of assessment - *Thoughts or plans indicating suicide risk* and *Relative / carer concern about suicide risk*. Where individuals had both a history of suicide attempts and current suicidal ideation, a category level “Both” was created.

#### Previous CDAT treatment

A binary variable indicated whether a service user had previously engaged in CDAT treatment prior to the index date. Although this study used the first available episode for each individual, this does not preclude those episodes which occurred prior to 2006 or in other boroughs which are not served by SLaM CDAT services.

#### Alcohol consumption (Drinks per Drinking Day)

Variation in severity of baseline alcohol use was captured through measures of quantity, from which a *Drinks per Drinking Day (DDD)* variable was derived, using a grouping previously utilised in a study of alcohol treatment outcomes in England using NDTMS data (37), as follows:

i. Abstinent (cases where either units per day or days drinking = 0);
ii. Low to high (1–15 units per drinking day);
iii. High to severe (16–30 units per drinking day);
iv. Extreme (≥31 units per drinking day).

This grouping also reflects the cut-offs used in NICE alcohol treatment guidelines as part of the process of establishing the appropriate level of treatment provision for a service user based on their alcohol use on treatment admission (38). Variation in frequency of drinking was not incorporated as the overwhelming majority of service users drank daily.

#### Non-alcohol drug use

Separate binary variables were derived for cocaine use, cannabis use, and other (non-alcohol, on-opioid) drug use.

#### Parental status

Parental status incorporated three levels: “Not a parent”, “Parent: no children living with service user” and “Parent: some or all children living with service user”.

#### Age

Age was derived from the month and year of birth and the treatment start date. Testing the linearity of *Age* as a continuous variable revealed some non-linearity when fit in its raw, centred, log and square root transformed forms, and so was consequently converted into a categorical variable with levels representing age groups 18-30, 31-40, 41-50 and over 51.

#### Sex

*Sex* was dichotomised as Male / Female.

#### Ethnicity

Ethnicity categories were collapsed into “White”, “Black”, “Asian”, “Mixed”, and “Other” for descriptive statistics. Due to low frequencies in the latter three categories, these were further collapsed into “White”, “Black”, and “Other” for the regression analysis.

#### Crisis care in year prior to CDAT treatment start

A binary variable was created to indicate any previous episode of care that involved psychiatric inpatient or community crisis care from the year prior to the index date.

#### Criminal Justice System involvement

A binary variable was created to indicate any self-reported involvement in criminal activity, and/or the treatment episode included an offender management programme, i.e., “Drug Treatment and Testing Order”, “Drug Rehabilitation Requirement” or “Drug Intervention Programme”.

#### Mental health problems

A binary variable was created to indicate mental illness or poor mental health (the latter identified by a TOP *Psychological health status* rating in the lower tertile).

#### Physical health problems

A binary variable was created to indicate physical illness or poor physical health (the latter identified by a TOP *Physical health status rating* in the lower tertile).

#### Social isolation

Social isolation was identified by the BRS-A item of the same name (a binary variable).

#### Housing status

A binary variable was created to indicate housing status (“Stable” vs “Homeless or Unstable”).

#### Other risk factors measured

Two other variables were extracted but omitted from the regression analysis due to high levels of missing data; *Quality of life* and *Employment status*. Both variables are reported in the summary statistics.

### Outcome: Crisis care contact or death by suicide

Due to death by suicide being a rare event, a composite binary variable was created denoting whether an event related to suicidal behaviour—either fatal or non-fatal—occurred in the year after commencing CDAT treatment. This variable was coded “Yes” if either of two qualifying events occurred in the year post-index, i.e., either death by suicide, or contact with psychiatric crisis care services. Deaths by suicide were identified by ICD-10 codes *X60 - X84 Intentional Self-Harm*, *Y10 - Y34 Event of undetermined intent*, *Y87.0 Sequelae of intentional self-harm* as underlying cause of death. This is in accord with the convention used by the ONS and the National Confidential Inquiry into Suicide and Safety in Mental Health (39). Time in days between index date and death were derived from the CRIS field *Date of death.* Crisis care contact events were identified by searching CRIS for any psychiatric inpatient or community crisis care episodes within the year after starting CDAT treatment, including contacts with Crisis Resolution Teams, the health-based Place of Safety (which receives individuals detained under the Mental Health Act), and liaison psychiatry teams (which assess individuals in psychiatric crisis in acute hospital emergency departments). The date of said contact was used to derive the time in days between index and crisis contact.

### Analysis

All statistical analyses were conducted in R version 4.2.1 (40). Cohort characteristics at baseline were described using frequencies, percentages, means and standard deviations as appropriate, and stratified by the outcome *Crisis care contact or death by suicide*.

A complete-case survival analysis was conducted using multivariable Cox proportional hazards regression to model the time-to-event, with the event being *Crisis care contact or death by suicide,* and underlying timescale being days since CDAT treatment start. Time-to-event was derived from the first occurring qualifying event, such that if an observation had a recorded crisis care contact event before later death by suicide or later re-contact with crisis care, the time-to-event was defined as the time to first crisis contact, with that observation remaining in a ‘failure’ state afterwards. Death by non-suicide cause was treated as a competing risk, as it precludes the occurrence of the event of interest. Observations were censored at the time of death by non-suicide cause (competing risk), or at 365 days after the index date if neither qualifying event or competing risk had occurred. Estimates of the increase or decrease in the instantaneous event probability at a given time associated with each risk factor are reported as Hazard Ratios (HR), 95% confidence intervals and p-values. Visual examination and tests of Schoenfeld residuals revealed that the proportional hazards assumption was not met for *Past-year crisis care,* and so a stratified Cox proportional hazards model was fit, i.e., a separate baseline hazard function was fit for both levels of *Past-year crisis care* (41).

#### Sensitivity analyses

##### Missing suicide risk screen

Cases excluded from the primary analysis due to missing data pertaining to history of suicide attempts and current suicidal ideation/concern were returned to the sample and coded as not having historic or current suicide risk identified (under the assumption that the missing data were not Missing At Random, and likely represented low-risk service users for whom the absence of a risk assessment indicates the absence of clinical concern). The analysis was repeated with this expanded data set and results compared.

##### Independent censoring in competing risks

In order to test the impact of non-independent censoring in the presence of competing risks, the Cox regression model was re-fit with the observations which were censored due to competing risk, i.e. non-suicide death, as i) censored at the end of the observation period with maximum possible follow-up time of 365 days reached and then ii) events (keeping follow-up time the same as in the primary analysis). Whilst this does not provide insight into whether the independent censoring assumption was violated in the primary analysis, it demonstrates the impact that the two extremes of non-independent censoring would have on effect estimates (41).

## Results

### Sample characteristics

Among 4451 CDAT alcohol treatment service users, the majority were male (68.7%, n=3060) and of white ethnicity (73.2%, n=3257), with a mean age of 41.6 years (SD=11.3). A history of suicide attempt was recorded in 31.5% (n=1403), and 18.4% (n=818) were recorded as either expressing current suicidal intent themselves or had friends or family expressing concern about their suicidal intent. Median units drunk per drinking day was 18; mean 20.6 (SD=16.4). See **Table 1** for sample characteristics.

**Table 1.** Descriptive statistics for cohort (n=4451) Stratified by outcome event: Crisis care contact or death by suicide within year of commencing CDAT treatment. Displayed as mean (M) and standard deviation (SD), or number (N) and percentage (%). P-values derived from chi-squared tests or t-test as appropriate to variable type.

| Variable | Range | M | SD | Event: No<br>M SD |  | Event: Yes<br>M SD |  | p-value |
| --- | --- | --- | --- | --- | --- | --- | --- | --- |
| Age | 18-86 | 41.6 | 11.3 | 41.8 | 11.3 | 40.5 | 11.0 | 0.025 |
| Variable | Categories | N | % | Event: No<br>n % |  | Event: Yes<br>n % |  | p-value |
| Gender | Female | 1391 | 31.3 | 1206 | 30.3 | 185 | 38.9 | <0.001 |
|  | Male | 3060 | 68.7 | 2770 | 69.7 | 290 | 61.1 |  |
| Ethnicity | White | 3257 | 73.2 | 2874 | 72.3 | 383 | 80.6 | 0.001 |
|  | Black | 690 | 15.5 | 643 | 16.2 | 47 | 9.9 |  |
|  | Asian | 189 | 4.2 | 167 | 4.2 | 22 | 4.6 |  |
|  | Mixed race | 156 | 3.5 | 145 | 3.6 | 11 | 2.3 |  |
|  | Other | 147 | 3.3 | 135 | 3.4 | 12 | 2.5 |  |
|  | Missing | 12 | 0.3 | 12 | 0.3 | 0 | 0.0 |  |
| Previously treated (CDAT) | Yes | 1222 | 27.5 | 1063 | 26.7 | 159 | 33.5 | 0.002 |
|  | No | 3102 | 69.7 | 2799 | 70.4 | 303 | 63.8 |  |
|  | Missing | 127 | 2.9 | 114 | 2.9 | 13 | 2.7 |  |
| Past-year psychiatric crisis care | Yes | 615 | 13.8 | 410 | 10.3 | 205 | 43.2 | <0.001 |
|  | No | 3836 | 86.2 | 3566 | 89.7 | 270 | 56.8 |  |
| Cocaine use | Yes | 894 | 20.1 | 804 | 20.2 | 90 | 18.9 | 0.598 |
|  | No | 3499 | 78.6 | 3123 | 78.5 | 376 | 79.2 |  |
|  | Missing | 58 | 1.3 | 49 | 1.2 | <10 | - |  |
| Cannabis use | Yes | 949 | 21.3 | 850 | 21.4 | 99 | 20.8 | 0.871 |
|  | No | 3445 | 77.4 | 3077 | 77.4 | 368 | 77.5 |  |
|  | Missing | 57 | 1.3 | 49 | 1.2 | <10 | - |  |

| Variable | Categories | N | % | Event: No |  | Event: Yes |  | p-value |
| --- | --- | --- | --- | --- | --- | --- | --- | --- |
|  |  |  |  | n | % | n | % |  |
| Other drug use | Yes | 287 | 6.4 | 244 | 6.1 | 43 | 9.1 | 0.017 |
|  | No | 4105 | 92.2 | 3682 | 92.6 | 423 | 89.1 |  |
|  | Missing | 59 | 1.3 | 50 | 1.3 | <10 | - |  |
| Alcohol DDD | Abstinent | 262 | 5.9 | 241 | 6.1 | 21 | 4.4 | <0.001 |
|  | Low to high | 1568 | 35.2 | 1441 | 36.2 | 127 | 26.7 |  |
|  | High to severe | 1857 | 41.7 | 1641 | 41.3 | 216 | 45.5 |  |
|  | Extreme | 707 | 15.9 | 603 | 15.2 | 104 | 21.9 |  |
|  | Missing | 57 | 1.3 | 50 | 1.3 | <10 | - |  |
| Suicide risk (combined) | Both | 436 | 9.8 | 335 | 8.4 | 101 | 21.3 | <0.001 |
|  | Current | 382 | 8.6 | 334 | 8.4 | 48 | 10.1 |  |
|  | Historic | 967 | 21.7 | 789 | 19.8 | 178 | 37.5 |  |
|  | No | 2666 | 59.9 | 2518 | 63.3 | 148 | 31.2 |  |
| Suicide risk: current plans or carer concern | Yes | 818 | 18.4 | 669 | 16.8 | 149 | 31.4 | <0.001 |
|  | No | 3633 | 81.6 | 3307 | 83.2 | 326 | 68.6 |  |
| History of suicide attempt | Yes | 1403 | 31.5 | 1124 | 28.3 | 279 | 58.7 | <0.001 |
|  | No | 3048 | 68.5 | 2852 | 71.7 | 196 | 41.3 |  |
| Physical health problems | Yes | 1503 | 33.8 | 1309 | 32.9 | 194 | 40.8 | 0.001 |
|  | No | 2948 | 66.2 | 2667 | 67.1 | 281 | 59.2 |  |
| Housing status | Stable | 3346 | 75.2 | 2999 | 75.4 | 347 | 73.1 | 0.282 |
|  | Unstable/Homeless | 1105 | 24.8 | 977 | 24.6 | 128 | 26.9 |  |
| Social isolation | Yes | 1327 | 29.8 | 1127 | 28.3 | 200 | 42.1 | <0.001 |
|  | No | 3123 | 70.2 | 2848 | 71.6 | 275 | 57.9 |  |
|  | Missing | <10 | - | <10 | - | <10 | - |  |
| Mental health problems | Yes | 2529 | 56.8 | 2148 | 54.0 | 381 | 80.2 | <0.001 |
|  | No | 1922 | 43.2 | 1828 | 46.0 | 94 | 19.8 |  |
| Quality of life (tertile) | 0-7 | 1198 | 26.9 | 1014 | 25.5 | 184 | 38.7 | <0.001 |
|  | 8-12 | 1313 | 29.5 | 1172 | 29.5 | 141 | 29.7 |  |
|  | 13-20 | 1037 | 23.3 | 984 | 24.7 | 53 | 11.2 |  |
|  |  |  |  | n | % | n | % |  |
|  | *Missing* | 903 | 20.3 | 806 | 20.3 | 97 | 20.4 |  |
| **Children** | No children | 2590 | 58.2 | 2310 | 58.1 | 280 | 58.9 | 0.002 |
|  | Children not living with service user | 984 | 22.1 | 860 | 21.6 | 124 | 26.1 |  |
|  | Children living with service user | 816 | 18.3 | 754 | 19.0 | 62 | 13.1 |  |
|  | *Missing* | 61 | 1.4 | 52 | 1.3 | <10 | - |  |
| **CJS involvement** | Yes | 555 | 12.5 | 497 | 12.5 | 58 | 12.2 | 0.915 |
|  | No | 3896 | 87.5 | 3479 | 87.5 | 417 | 87.8 |  |
| **Employment status** | In ETE | 1323 | 29.7 | 1238 | 31.1 | 85 | 17.9 | <0.001 |
|  | Economically inactive<sup>1</sup> | 236 | 5.3 | 203 | 5.1 | 33 | 6.9 |  |
|  | Unemployed | 2135 | 48.0 | 1855 | 46.7 | 280 | 58.9 |  |
|  | Other or unknown | 166 | 3.7 | 151 | 3.8 | 15 | 3.2 |  |
|  | *Missing* | 591 | 13.3 | 529 | 13.3 | 62 | 13.1 |  |

DDD = Drinks per Drinking Day, CJS = Criminal Justice System, ETE = Education, Training or Employment. <sup>1</sup>Economically inactive = Retired, or long-term sick or disabled. Frequencies <10 suppressed to prevent de-anonymisation

### Crisis care contact or death by suicide within year

There were 468 (10.5%) crisis care contact events within a year of starting CDAT treatment, and fewer than 10 deaths by suicide. Median time to crisis care contact was 95 days (range = 365 days). Median time to death by suicide was 160 days (range = 326 days). The majority of crisis care contacts were presentations to the Emergency Department (ED) (n=397, 84.8%), with 8.3% (n=39) accessing CRT services and 6.2% (n=31) accessing the Place of Safety. There were 49 non-suicide deaths which were not preceded by a crisis care contact event.

### Cox regression: Crisis care contact or death by suicide

There were n=4312 cases without any missing data across all covariates, and thus comprised the sample for the Cox proportional hazards model (n=139 [3.1%] removed due to missing data). There were 462 psychiatric crisis care contacts or deaths by suicide within this complete-case sample. The fully adjusted model was fit with all covariates included, and the competing risk—death by non-suicide cause—censored as per protocol described above. Unadjusted and fully adjusted hazard ratios, confidence intervals and p-values from the stratified model are shown in **Table 2**, with the fully adjusted model displayed in forest plot form in **Figure 2**.

**Figure 2.**
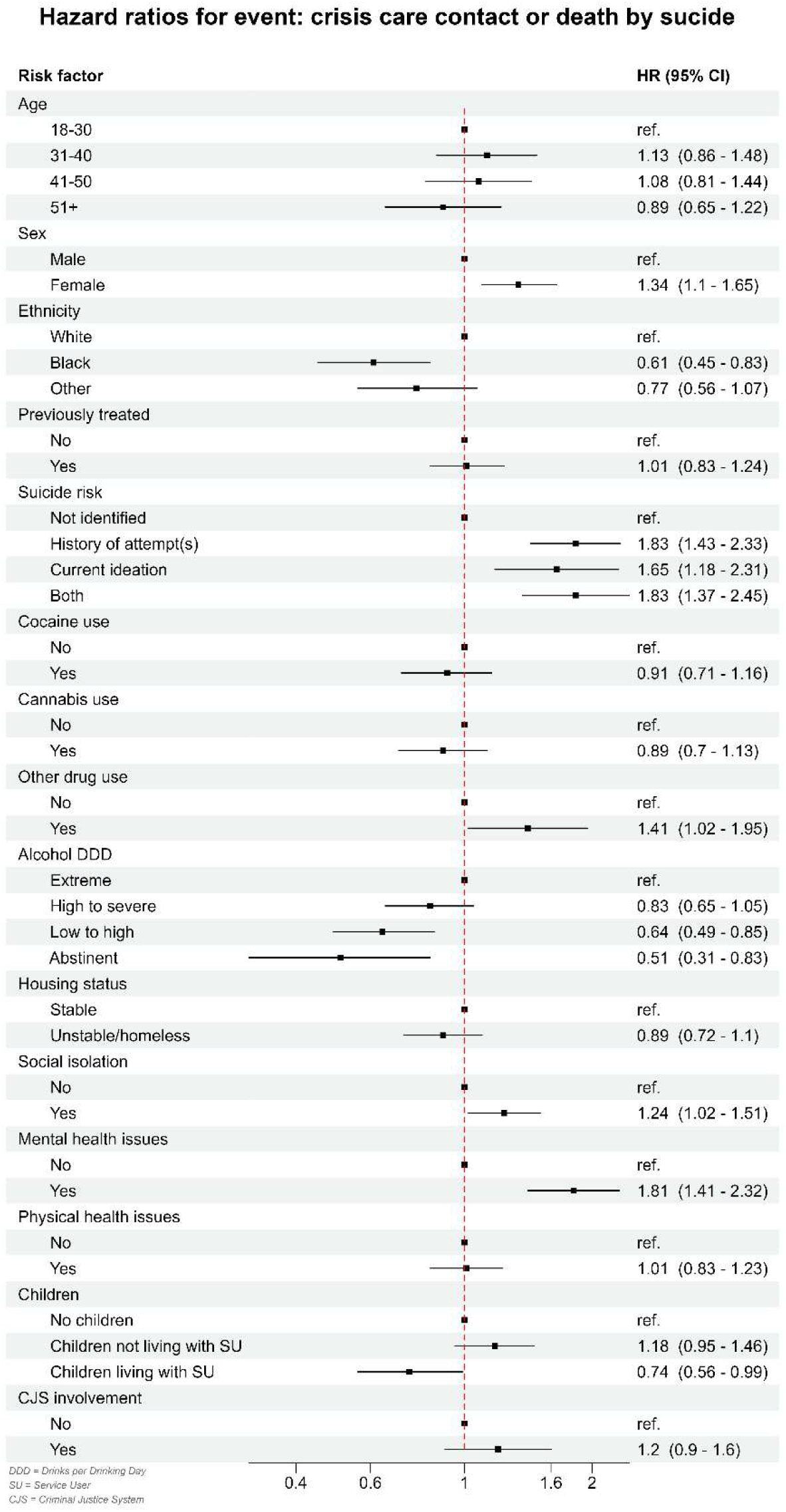
Forest plot of results from fully adjusted Cox proportional hazards regression Stratified by Past-year psychiatric crisis care Event = Crisis care contact or death by suicide within year of commencing CDAT treatment. n=4312. Events = 462

**Table 2.**
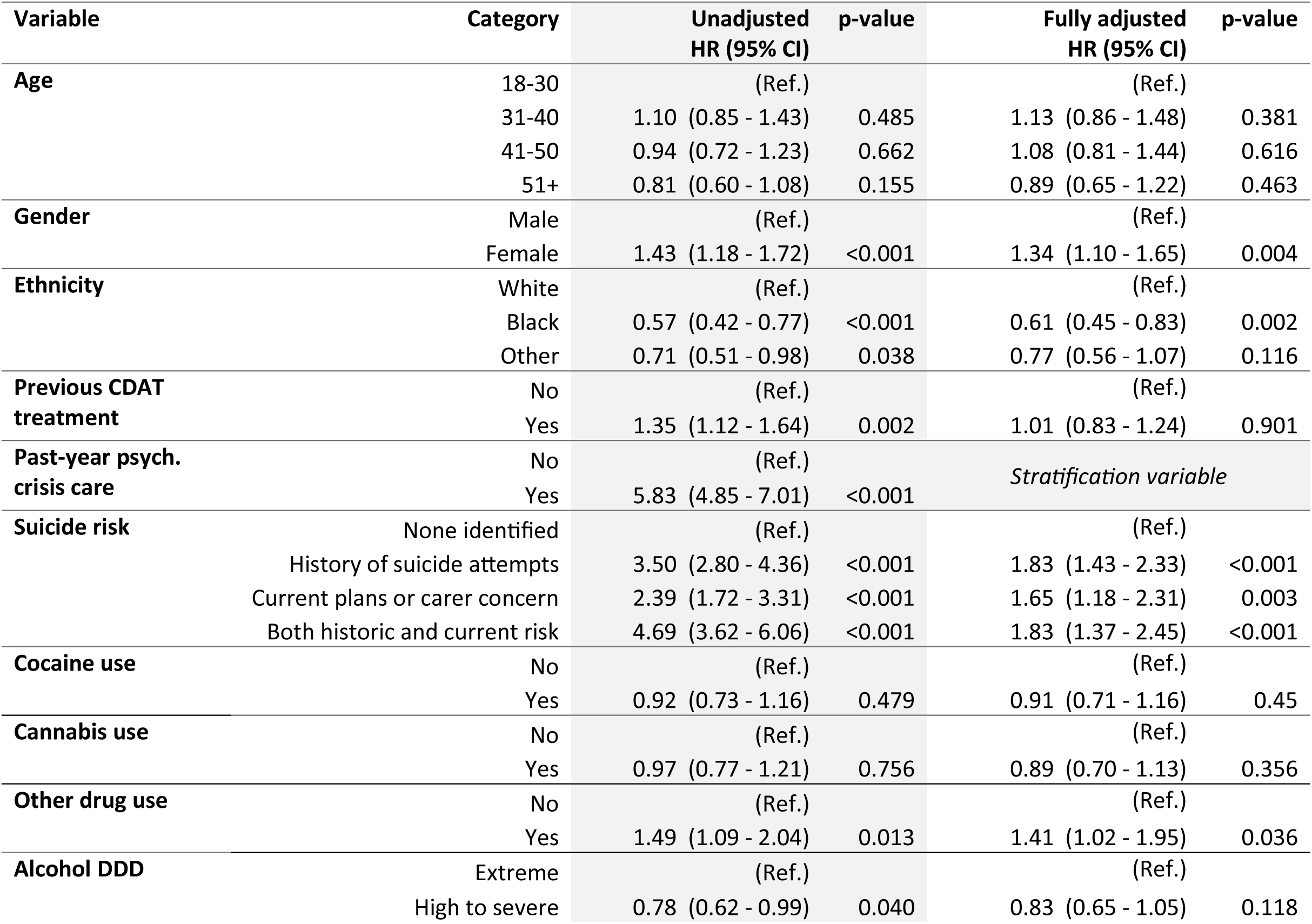
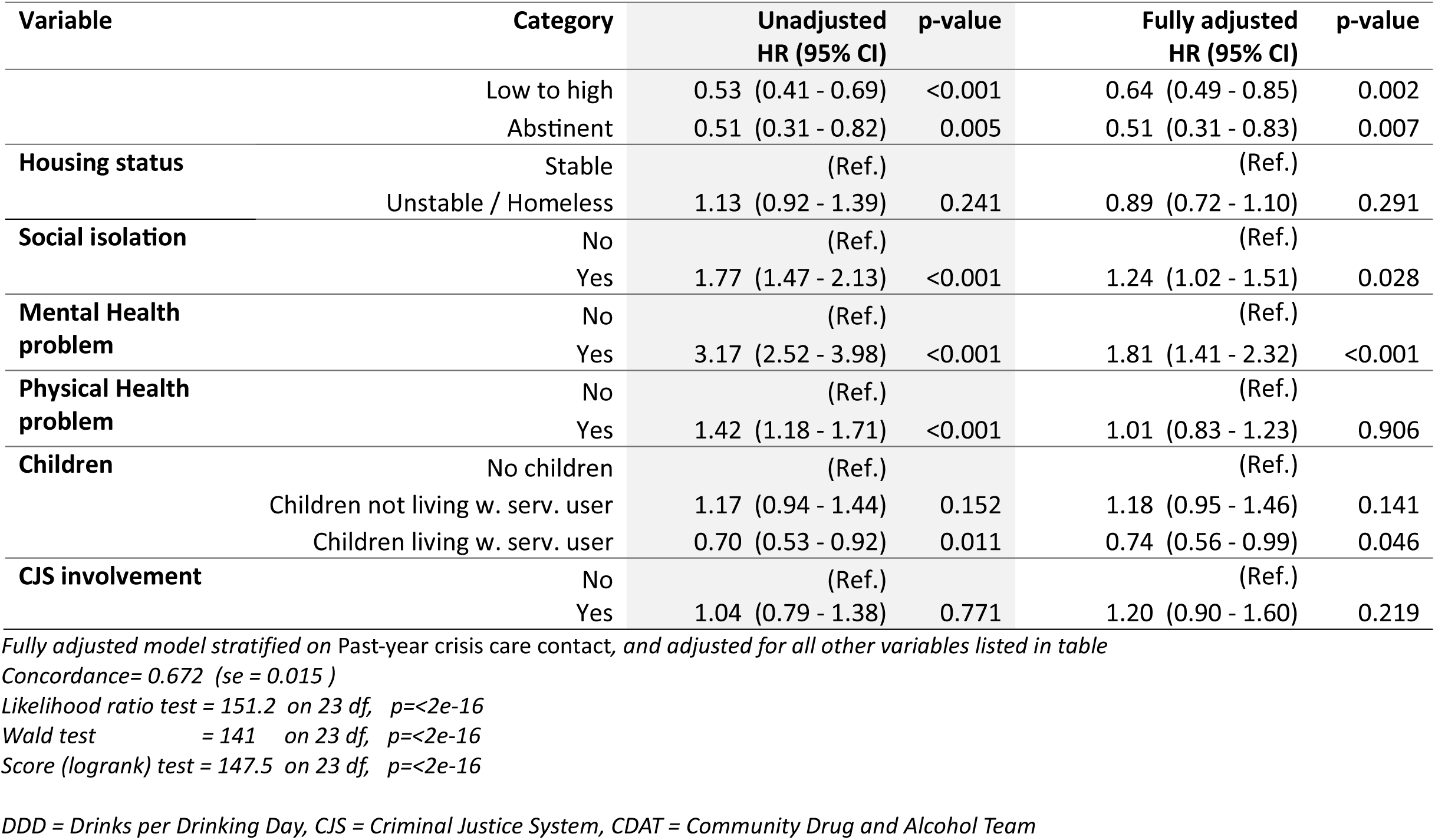
Cox proportional hazards regression. Event = Crisis care contact or death by suicide within one year of CDAT treatment commencement. n=4312. Events = 462

In the stratified and fully adjusted model, increased risk of crisis care contact or death by suicide within one year of CDAT treatment commencement was associated with; female sex (HR 1.34, 1.10-1.65); history of suicide attempt (HR 1.83, 1.43-2.33); current suicidal plans or carer concern at baseline (HR 1.65, 1.18-2.31); both history of suicide attempt and current suicidal plans / concern at baseline (HR 1.83, 1.37-2.45); other drug use, not including cocaine and cannabis (HR 1.41, 1.02-1.95); social isolation (HR 1.24, 1.02-1.51); and mental health problems (HR 1.81, 1.41-2.32). Decreased risk was associated with drinking 1-15 units of alcohol per drinking day (HR 0.64, 0.49-0.85, ref: >30 units); alcohol abstinence at baseline (HR 0.51, 0.31-0.83, ref: >30 units); black ethnicity (HR 0.61, 0.45-0.83); and children living with the service user (HR 0.74, 0.56-0.99).

### Sensitivity Analyses

The hazard ratio estimates did not deviate substantially from the primary analysis when fit to any of the hypothetical scenarios described above (see Methods). See **Supplementary Table 3** for hazard ratios after the observations with a missing suicide risk screen were returned to the sample. See **Supplementary Table 4** for hazard ratios under the simulated extremes of non-independent censoring in the presence of a competing risk (non-suicide death).

## Discussion

Among a sample of 4451 alcohol treatment service users, fewer than 10 died by suicide, and 10.5% came into contact with psychiatric crisis care, within the year following treatment start. After adjustment, a history of suicide attempt was associated with approximately 80% increased hazard of crisis care contact or death by suicide, and current suicidal ideation in the absence of historical attempt was associated with a 65% increased hazard of the same outcome. Increases in hazard of crisis care contact or death by suicide were also associated with mental health problems; use of drugs other than opioids, cocaine, cannabis or alcohol; female sex; and social isolation. Abstinence from alcohol at treatment start was associated with a 50% reduction hazard of crisis care contact or death by suicide (compared to drinking >30 units per drinking day). Decreases in hazard were also associated with drinking 1–15 units per drinking day (compared to drinking >30 units per drinking day); Black ethnicity compared to White; and children living with the service user compared to not being a parent.

### Findings in context

The increased hazard associated with a history of suicide attempt concurs with previous findings, both from addiction treatment settings (19,22,23,25,28) and general population studies (42) in which previous suicide attempt is considered one of the strongest predictors of future death by suicide.

Current suicidal ideation in the absence of an historic attempt was also associated with a significant increase in hazard of later crisis care contact or death by suicide, supporting results from crisis assessment settings that found that whilst acute alcohol use and binge drinking are associated with transient suicidal intent, alcohol dependence is not, and is the form of alcohol use most associated with later suicidal behaviour (43,44).

The decreased hazard of crisis care contact or death by suicide associated with black ethnicity is consistent with other UK research that found Black Caribbean and especially Black African ethnicity groups have lower suicide rates than White British groups (45), and a recent NCISH case-control study in which cases who died by suicide within 12 months of contact with a drug and alcohol service were less likely to be of an ethnic minority than non-suicide controls (8). Regarding sex, as the majority of events comprising the composite outcome in this study were non-fatal crisis contact events, our findings are consistent with three previous studies from addiction treatment-seeking populations which found female sex to be associated with double the odds of suicide attempt in the year following treatment start compared to male sex (28), but decreased hazard of death by suicide (20,26).

The decreased hazard among service users with children in their care is consist with the protective effect of parenthood found in general population samples (13). Whilst a previous study among CDAT service users found no difference in lifetime suicide attempt history between mothers and non-mothers, and between mothers with children under their care and mothers with children in alternative care, the study did not have any prospective follow-up after treatment start, so could not report whether the lifetime suicide attempts preceded or followed parenthood (46).

The increase in hazard of crisis care contact or death by suicide associated with social isolation is consistent with findings from other previous studies of addiction treatment cohorts: Darke et al. found social isolation to be associated with a four-fold increase in odds of suicide attempt within three years of starting substance use treatment (24); Pavarin et al. found the only social risk factor associated with death by suicide over 41 years of follow-up in Italy was being separated or divorced (26); and the NCISH found social isolation to feature in 14% of 100 serious incident reports on people who had died by suicide within recent contact with drug and alcohol services (8). Evidence from psychiatric patients has also found that loneliness in patients with substance misuse problems is particularly strongly associated with adverse outcomes (47).

Previous studies from cohorts of AUD patients accessing addiction treatment services have found mixed findings in terms of the effect of alcohol consumption pattern on suicide-related outcomes, though the effect of baseline levels of consumption appears to dissipate over time. For example, number of days of alcohol use at baseline has been found to be associated with increased odds of suicide attempt within 12 months (12), whereas maximum number of drinks in a 24-hr period was not associated with suicide attempt within 5 years (23).

The use of drugs other than cocaine, cannabis, opioids and alcohol was associated with an increased hazard of crisis care contact or death by suicide, however, no such association was found for cannabis use or cocaine use. These findings are consistent with those of the NCISH study of deaths by suicide in recent drug and alcohol service users, which found no difference between suicide decedent cases and matched living controls in terms of likelihood of cocaine or cannabis use, but did find an increased likelihood of use of other (non-opioid) drugs among suicide decedents (8).

### Clinical implications

The risk factors for suicidal behaviour among people accessing CDAT alcohol treatment identified in this study provide contextual knowledge that can aid patient-centred suicide assessment and safety planning in this population. They provide a potential framework which the service user and clinician can use in the process of exploring and establishing the individual service user’s constellation of risk factors relative to others in the same population, and identify potential avenues for intervention. Sorting risk factors according to those which are modifiable, and those which are protective, can be helpful in safety planning and identifying targets for intervention (see **Figure 3**, adapted from Hawton et al (2022) (9)). For example, for service users who are socially isolated, activities which involve connectedness may help modify this risk, such as engagement in mutual aid and peer support groups, or activities designed under the NHS social prescribing model (48).

**Figure 3.**
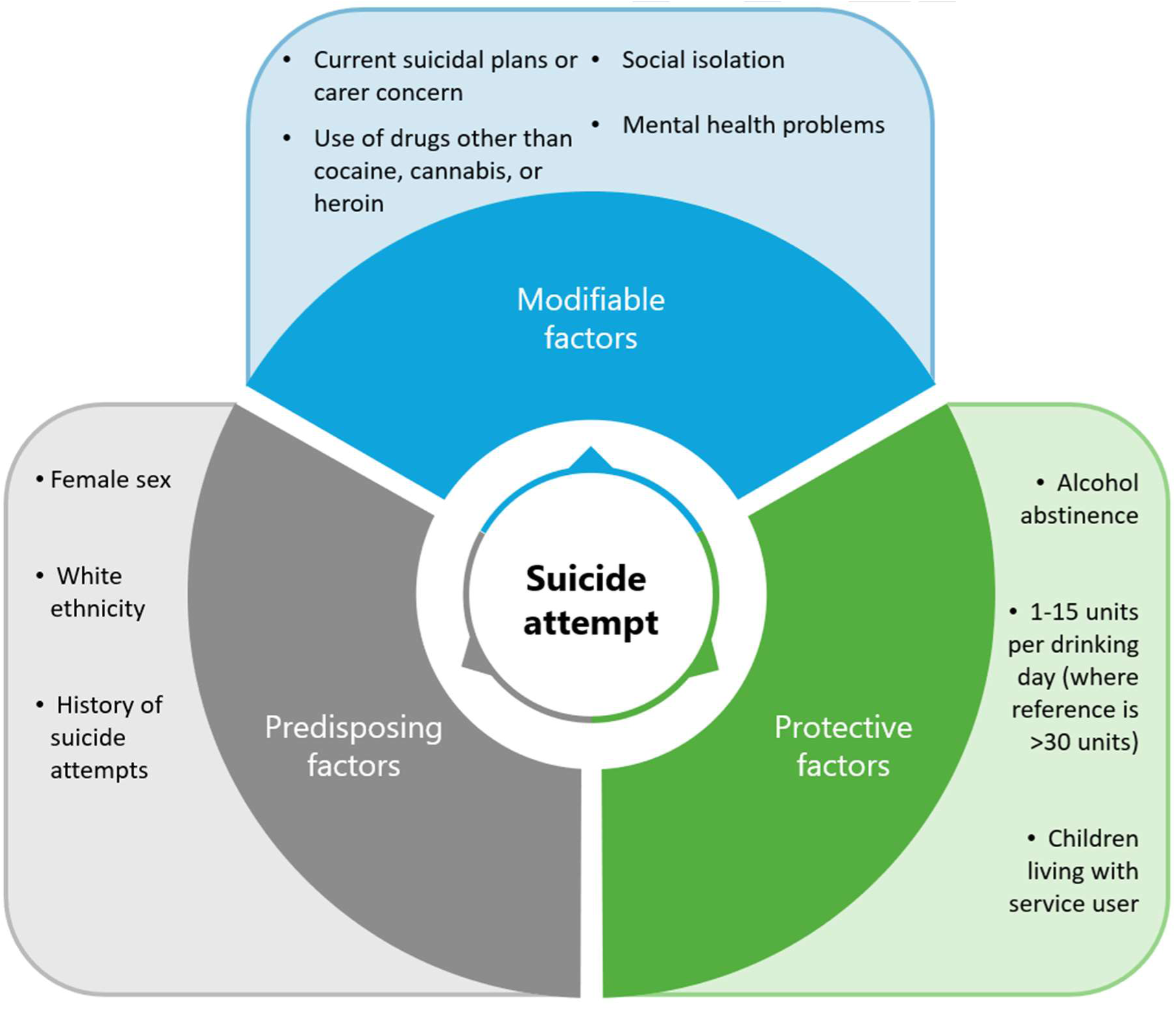
Risk factors for crisis care contact or death by suicide among CDAT alcohol treatment service users, within 1 year of starting treatment, as identified in this study.

The framework may also assist with staff training, as many staff working in drug and alcohol services receive little or no training in suicide risk factors (49).

### Strengths

This is the first prospective analysis of risk factors for suicidal behaviour in a purely alcohol-using sample accessing community-based addiction care. This population represent the largest proportion of CDAT service use, with a uniquely elevated suicide risk. The analysis uses 14 years’ of data with a rich array of structured fields used to define variables within tight window of inclusion (index +/- 14 days).

### Limitations

The composite outcome variable uses crisis care contact as a proxy for suicide attempt, and it is possible that not all such events were due to suicidal behaviour, e.g. psychotic symptoms related to stimulant use or alcohol withdrawal delirium. However, alcohol use is generally associated with suicidal rather than psychotic crisis presentations (50), and psychosis is itself a risk factor for a risk factor for suicidal behaviour (51).

Due to reporting restrictions around CRIS data, designed to prevent de-anonymisation, it is not possible to report the precise number of deaths by suicide beyond ‘fewer than ten’. A meta-analysis of 31 studies featuring 36,375 patients treated for AUD reported a proportional suicide mortality of 7.34% (5.7-8.98%) of all deaths (4), and the proportion of deaths that were by suicide in this cohort is within this expected band. Some deaths by suicide may not have been classified as such, especially as the majority of the study period occurred before the change in the ‘standard of proof’ required for a suicide conclusion to be reached at inquest (52). However, the definition of suicide used here is that used by coroners in the UK.

The risk factors identified in this study are not an exhaustive list; there will be some residual confounding from risk factors which could not be included due to sparse data (e.g. psychiatric diagnosis, which is not routinely recorded in NDTMS data), and the role of anticipated factors which may modify suicide risk after treatment start (e.g. relapse) are an avenue for further study.

### Conclusions

Understanding the demographic, circumstantial and clinical risk factors for suicide attempt among community alcohol treatment service users is a priority for suicide prevention. Patient-centred risk formulation and safety planning should include discussion of the impact of a range of risk factors including social isolation, alcohol consumption, other drug use, mental health and family circumstances.

## Data Availability

The ethical approval to access CRIS data (Oxfordshire Research Ethics Committee C (18/SC/0372)) requires the data to be stored behind an NHS firewall with access governed by a patient-led oversight committee. For this reason, the data cannot be made available in the manuscript, Supporting Information files or a public repository. However, subject to approval from the oversight committee, data access for research purposes is encouraged. Further information is available from.

## Supplementary Material

**Supplementary Table 1.**
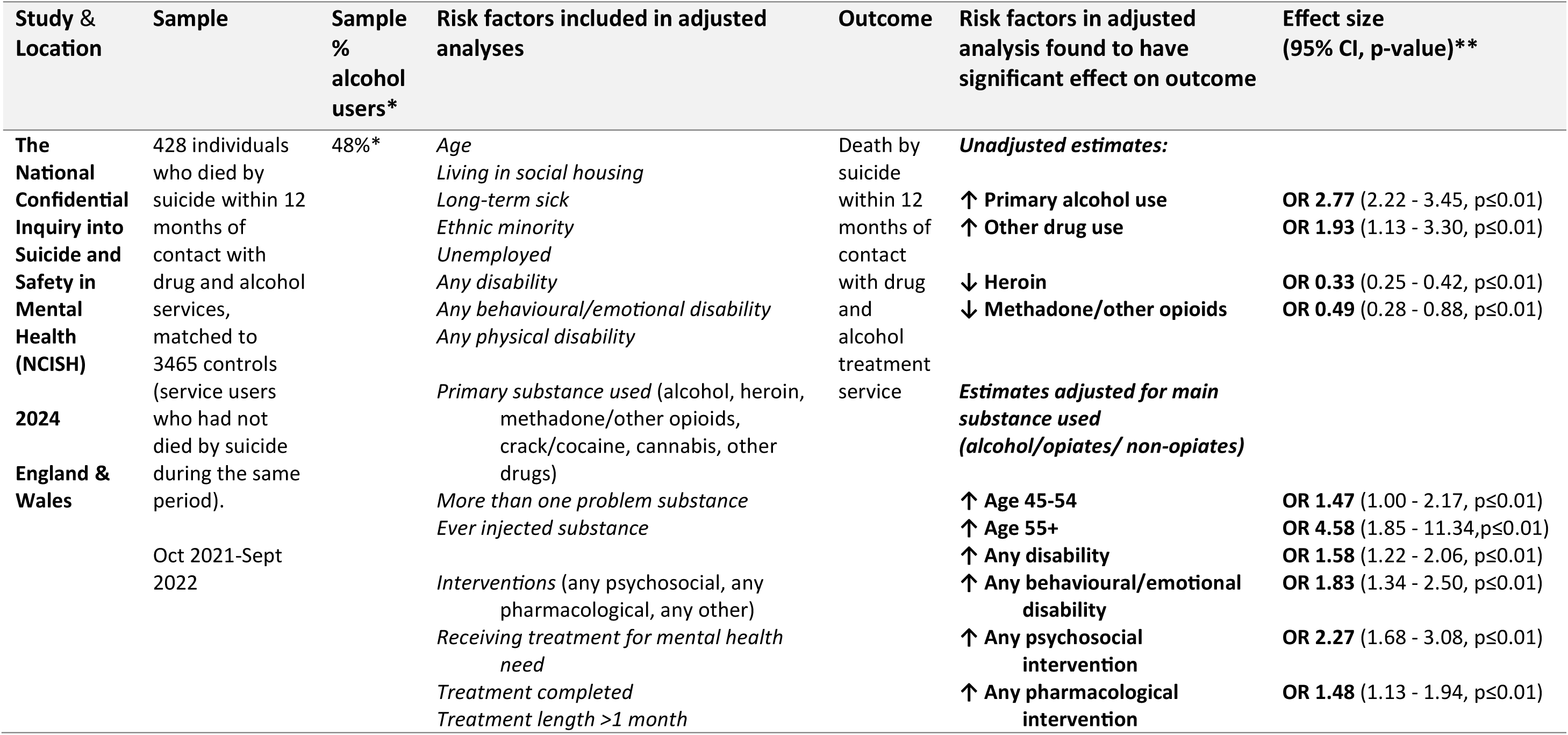
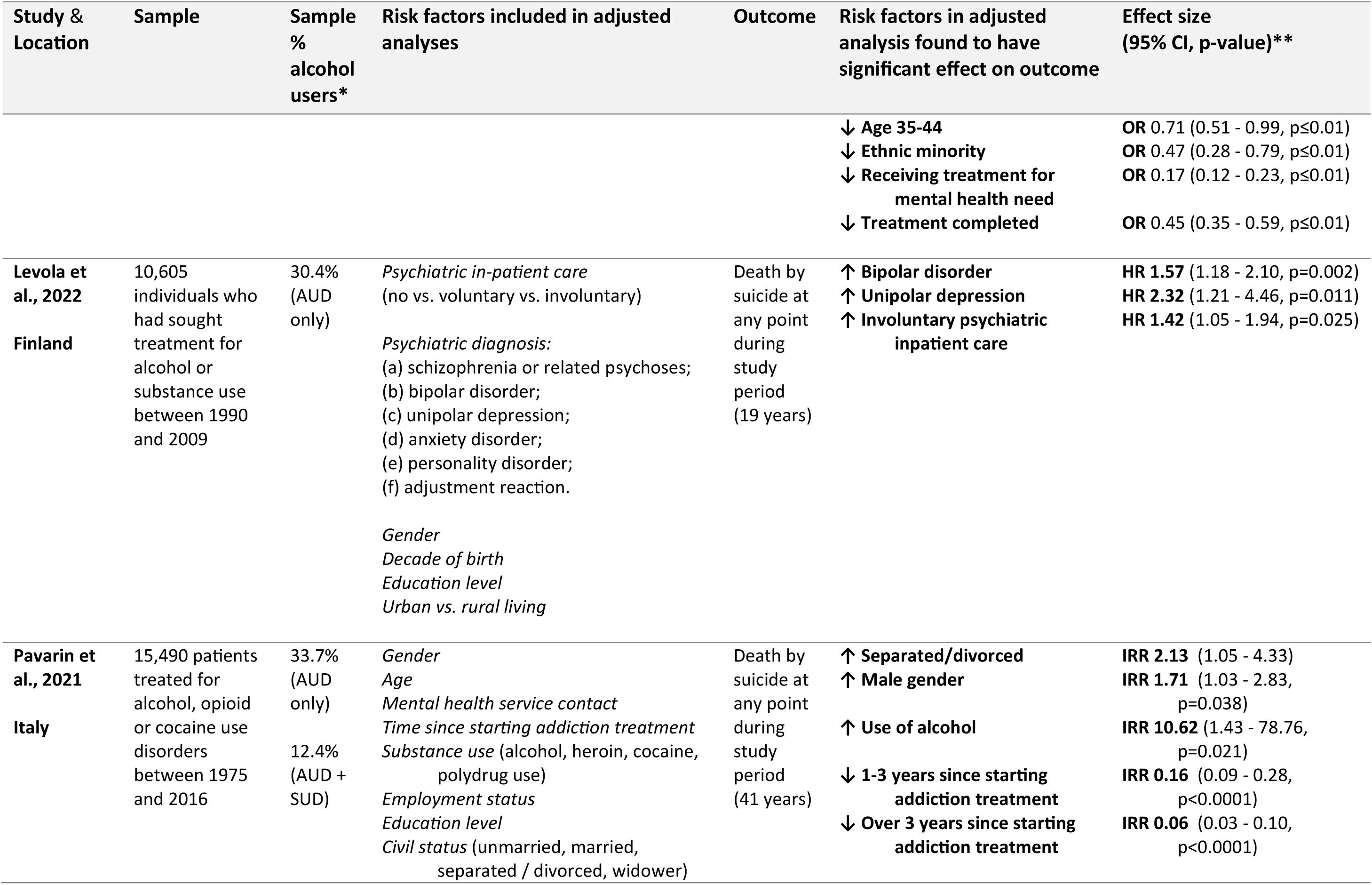
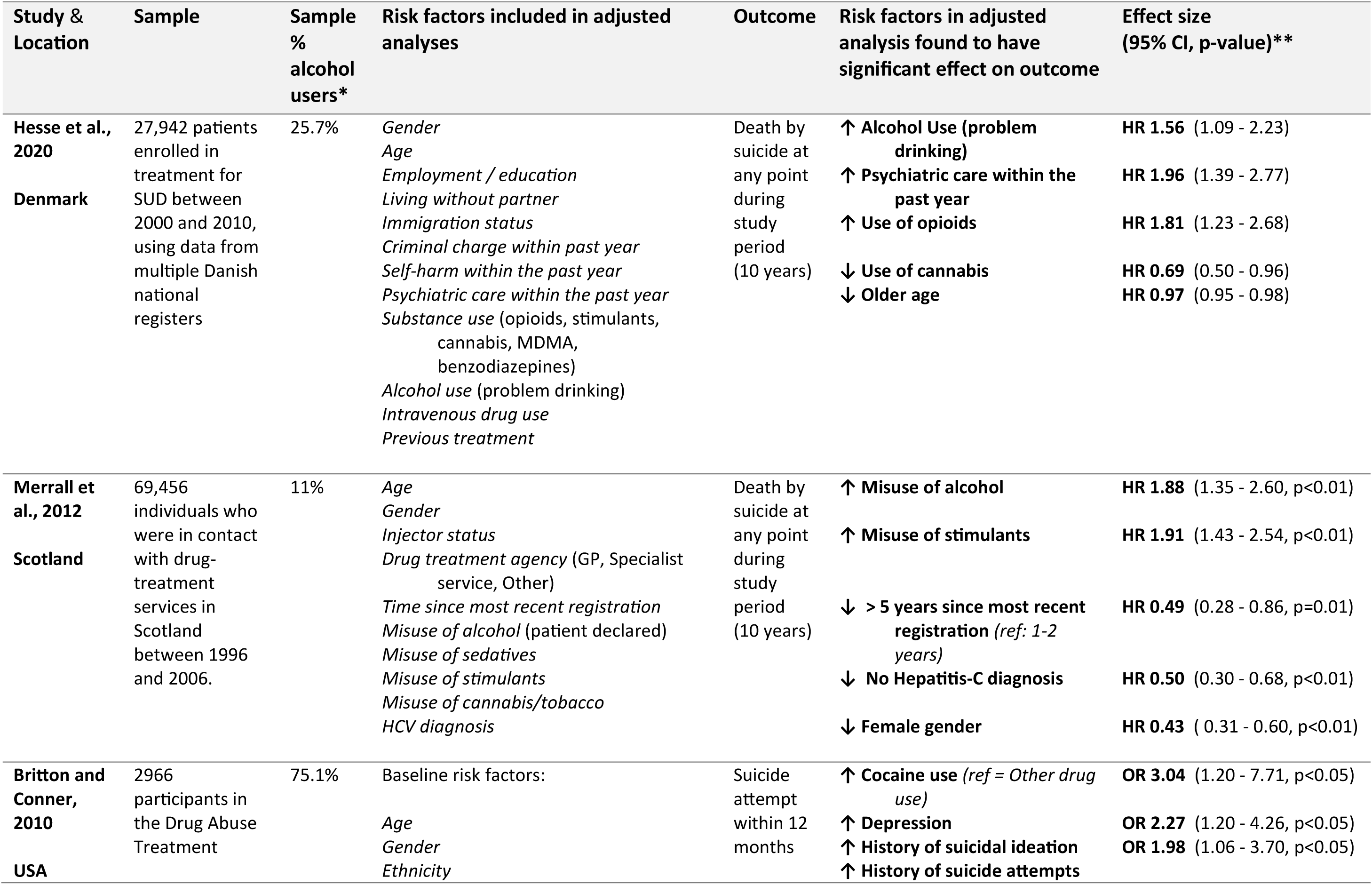
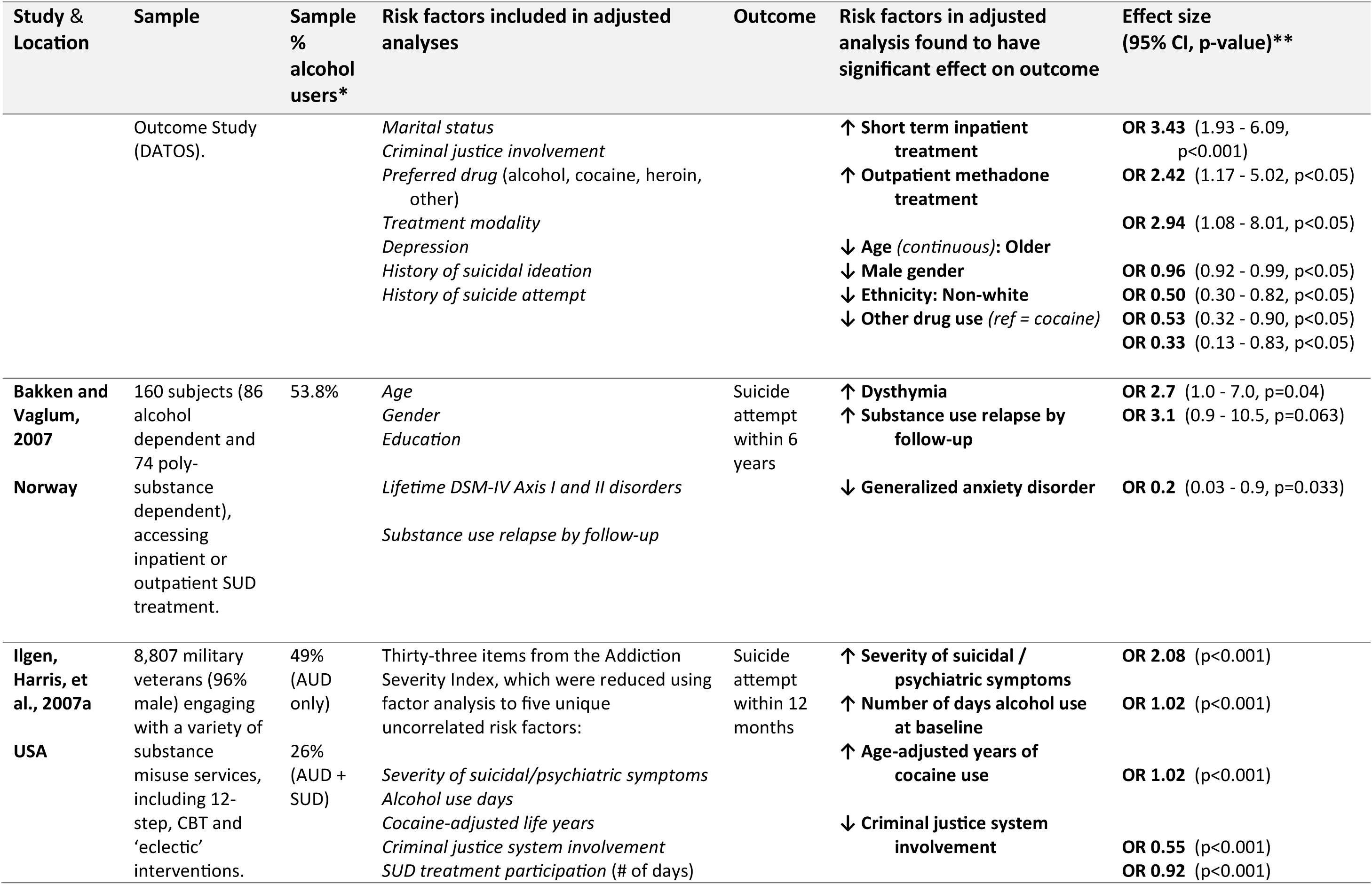
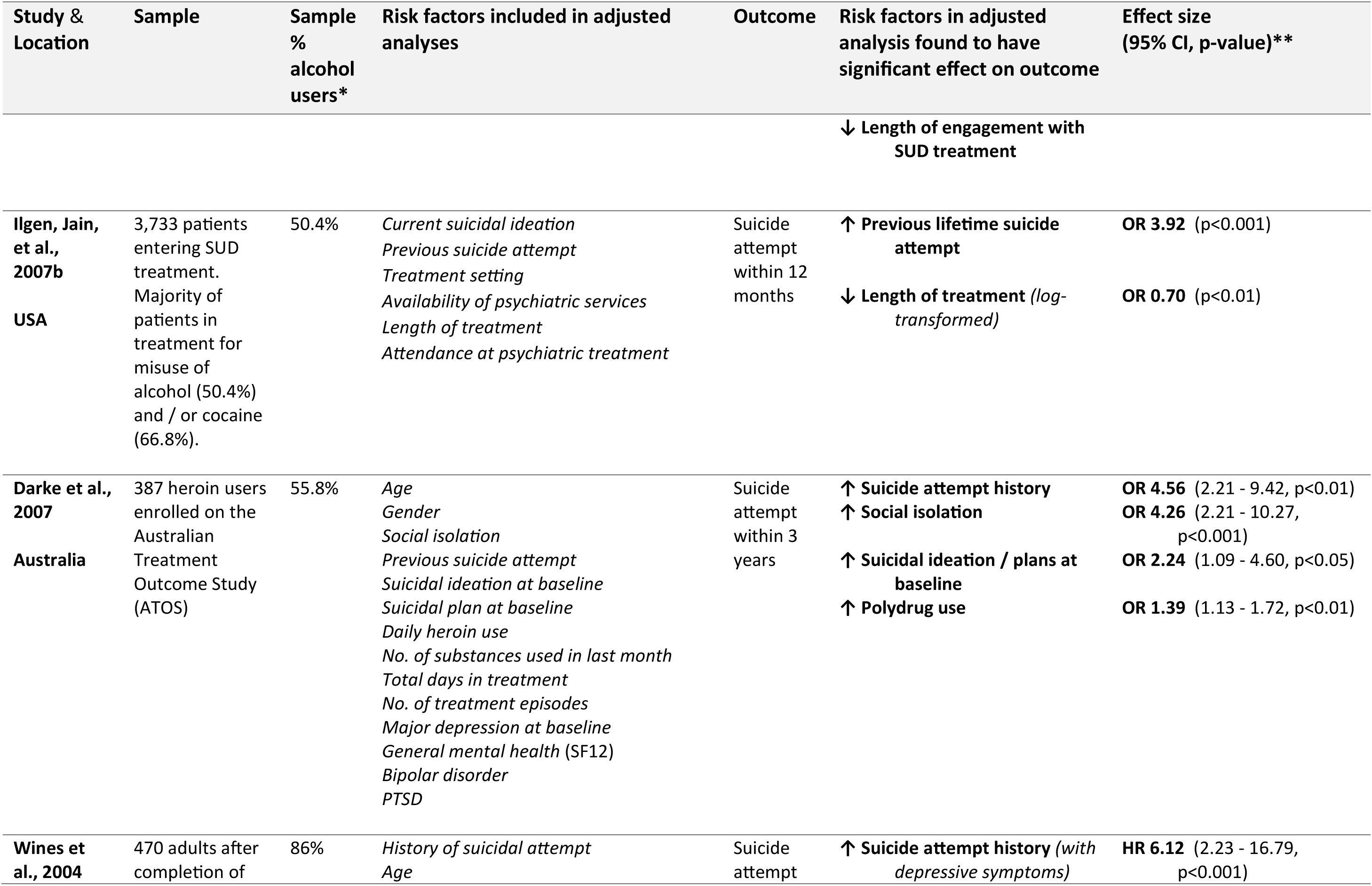
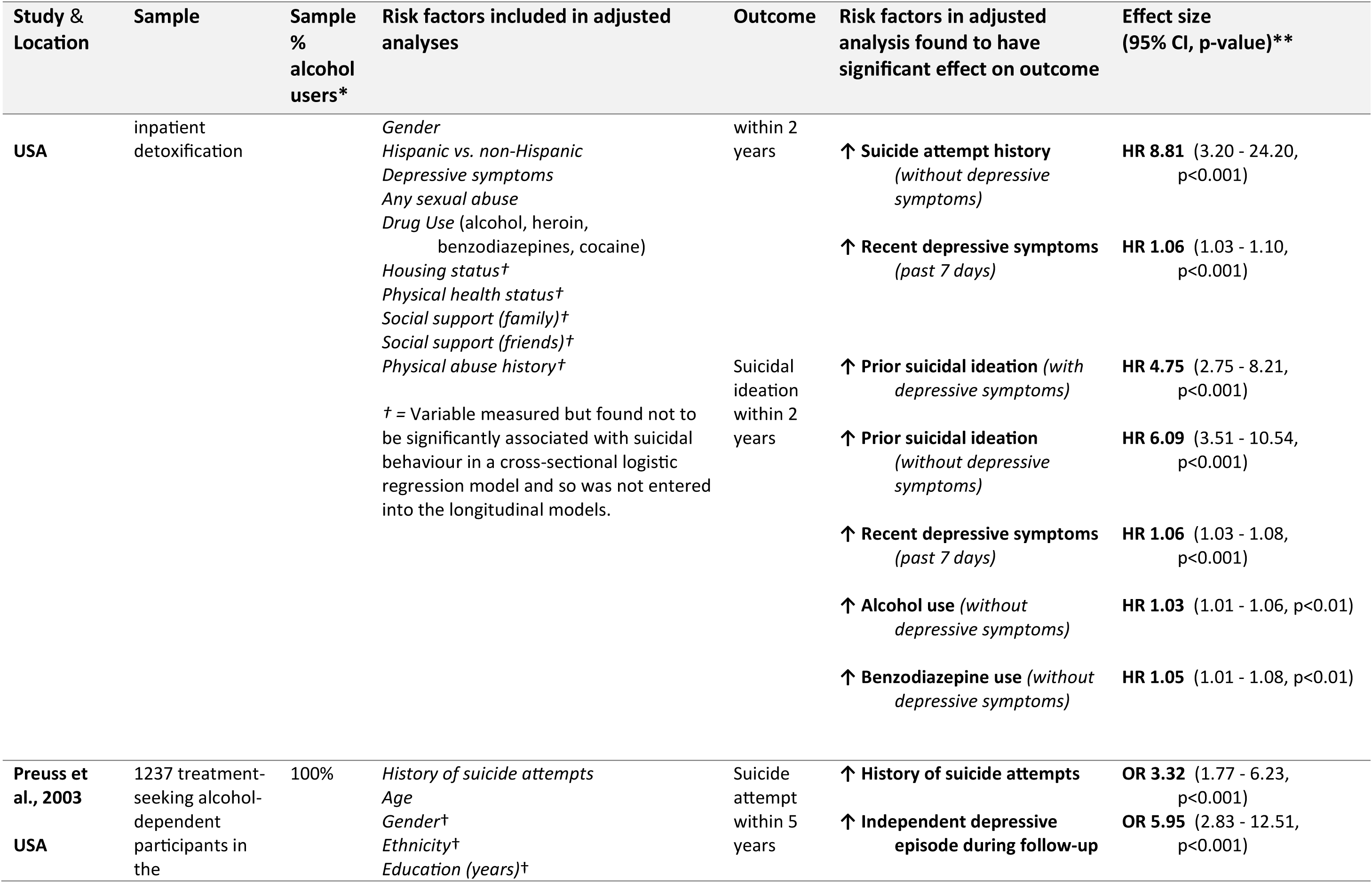
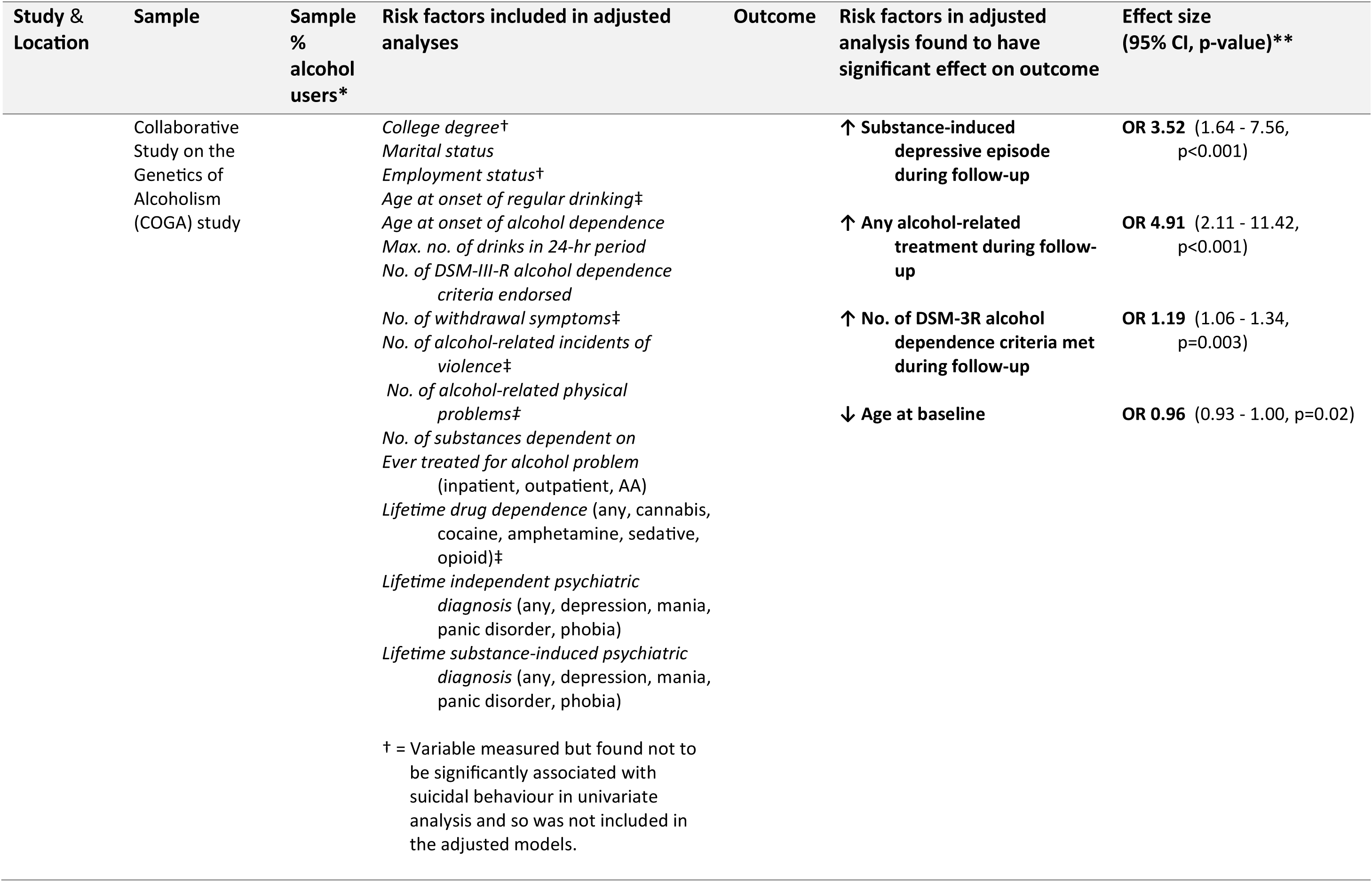
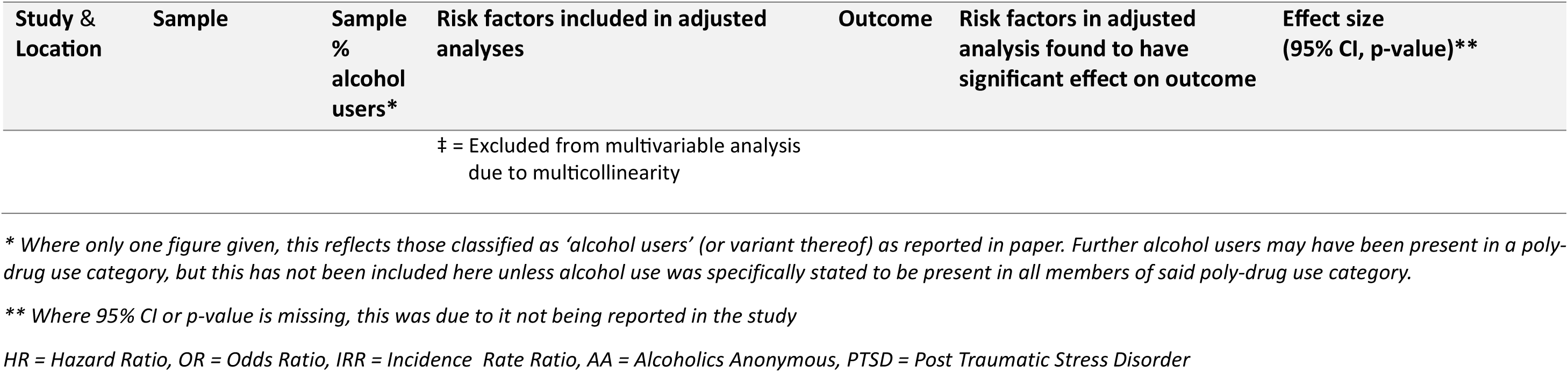
Summary of risk factors for suicidal behaviour identified by previous studies. All studies included below involve cohorts engaged with some form of SUD treatment involving alcohol at least to a minimal degree, and with a suicide-related outcome measured after treatment commencement.

**Supplementary Table 2.**
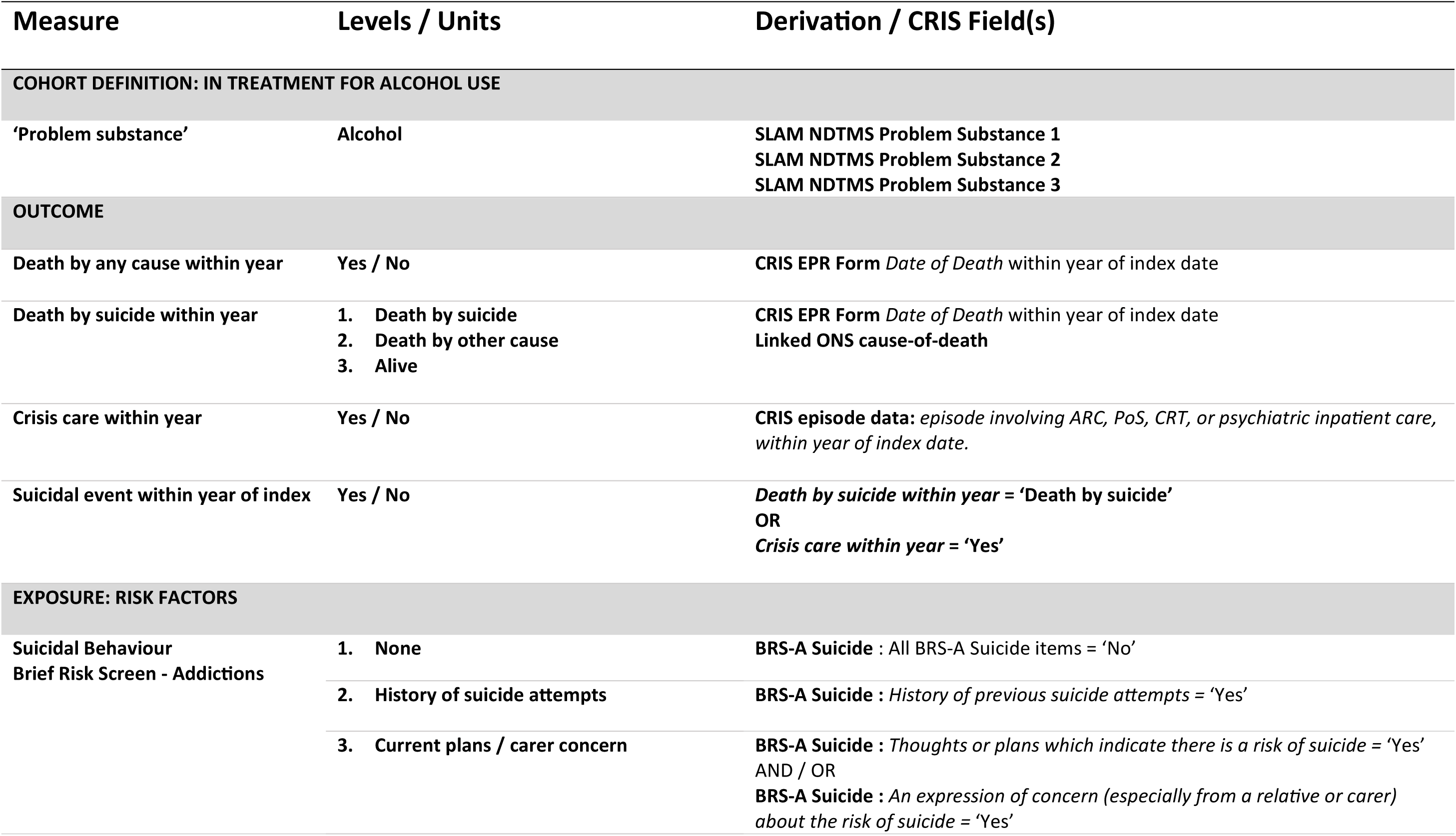
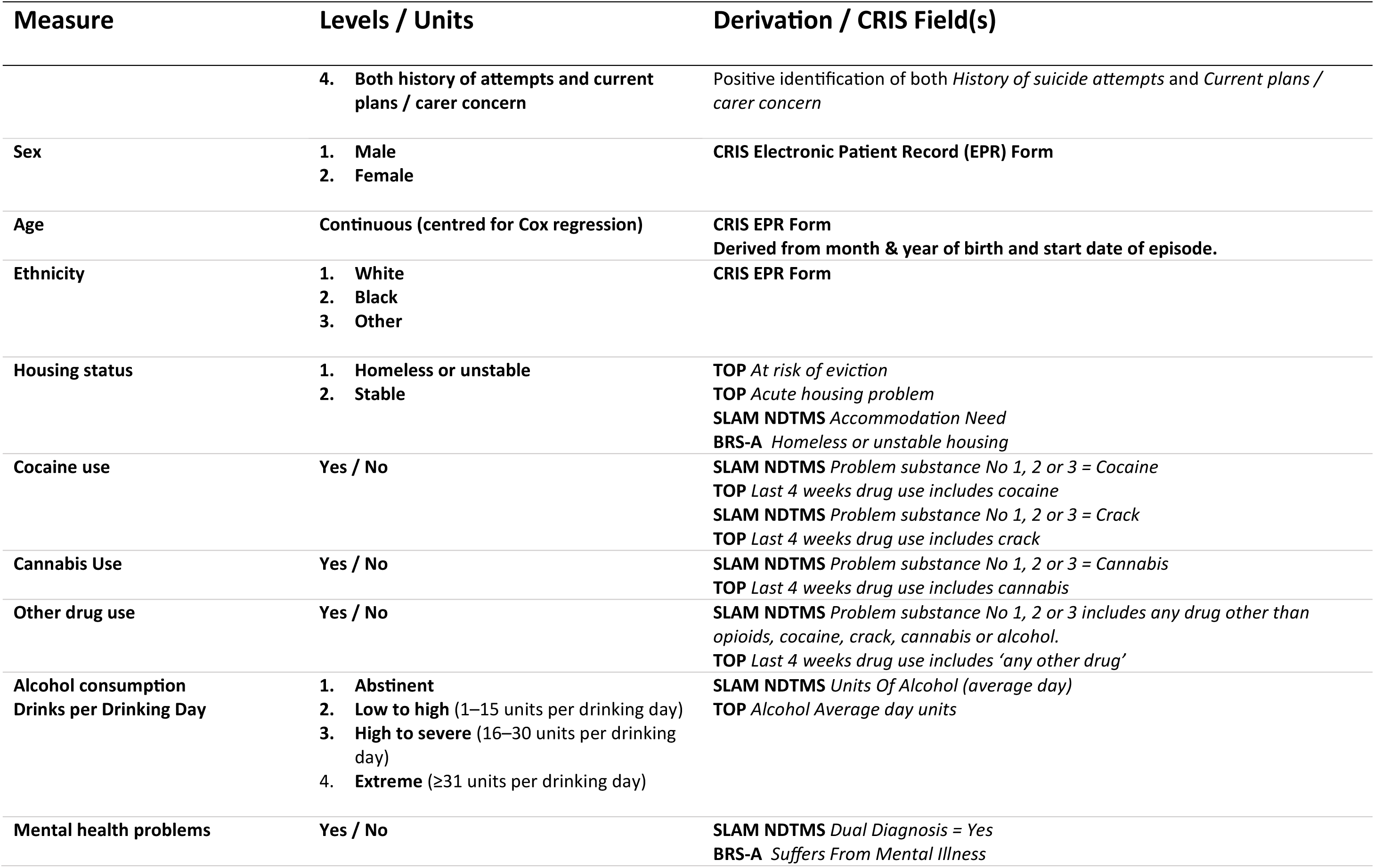
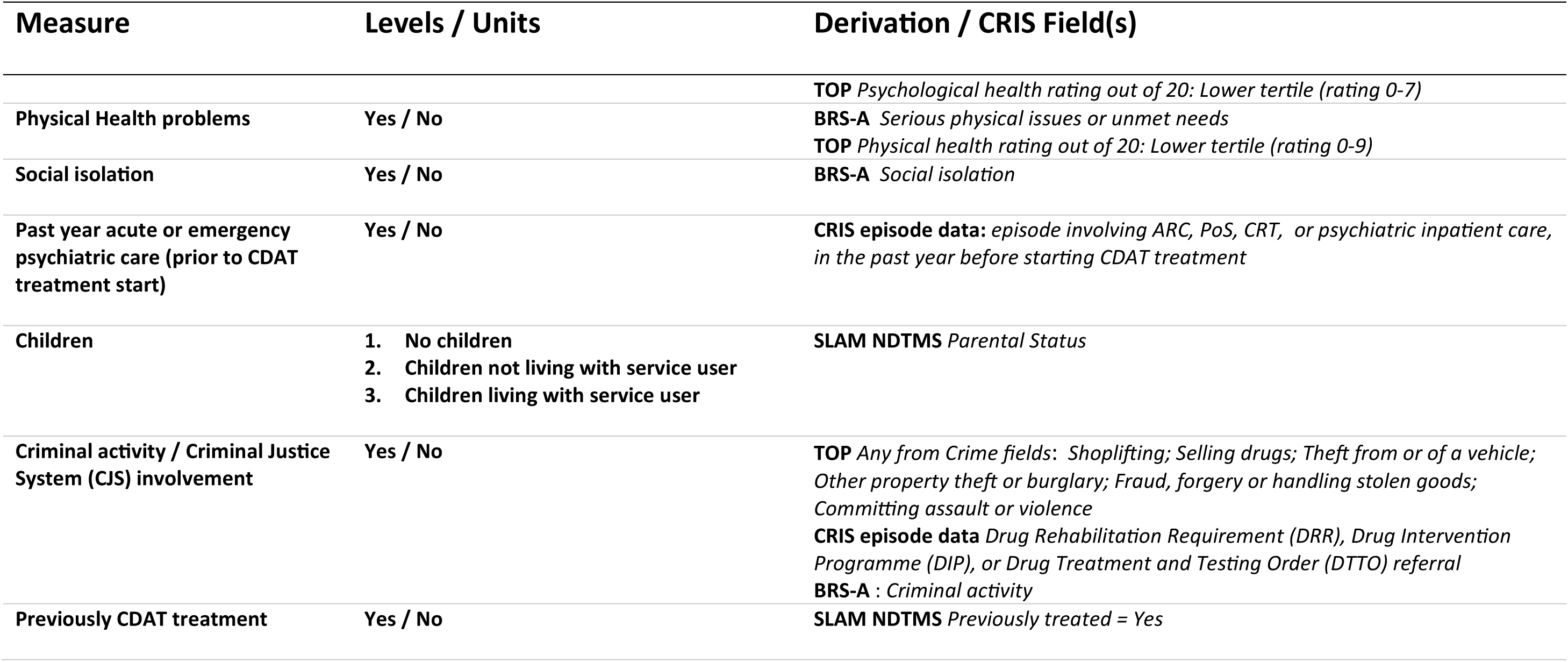
Summary of measures derived from electronic health records for analysis.

**Supplementary Table 3.**
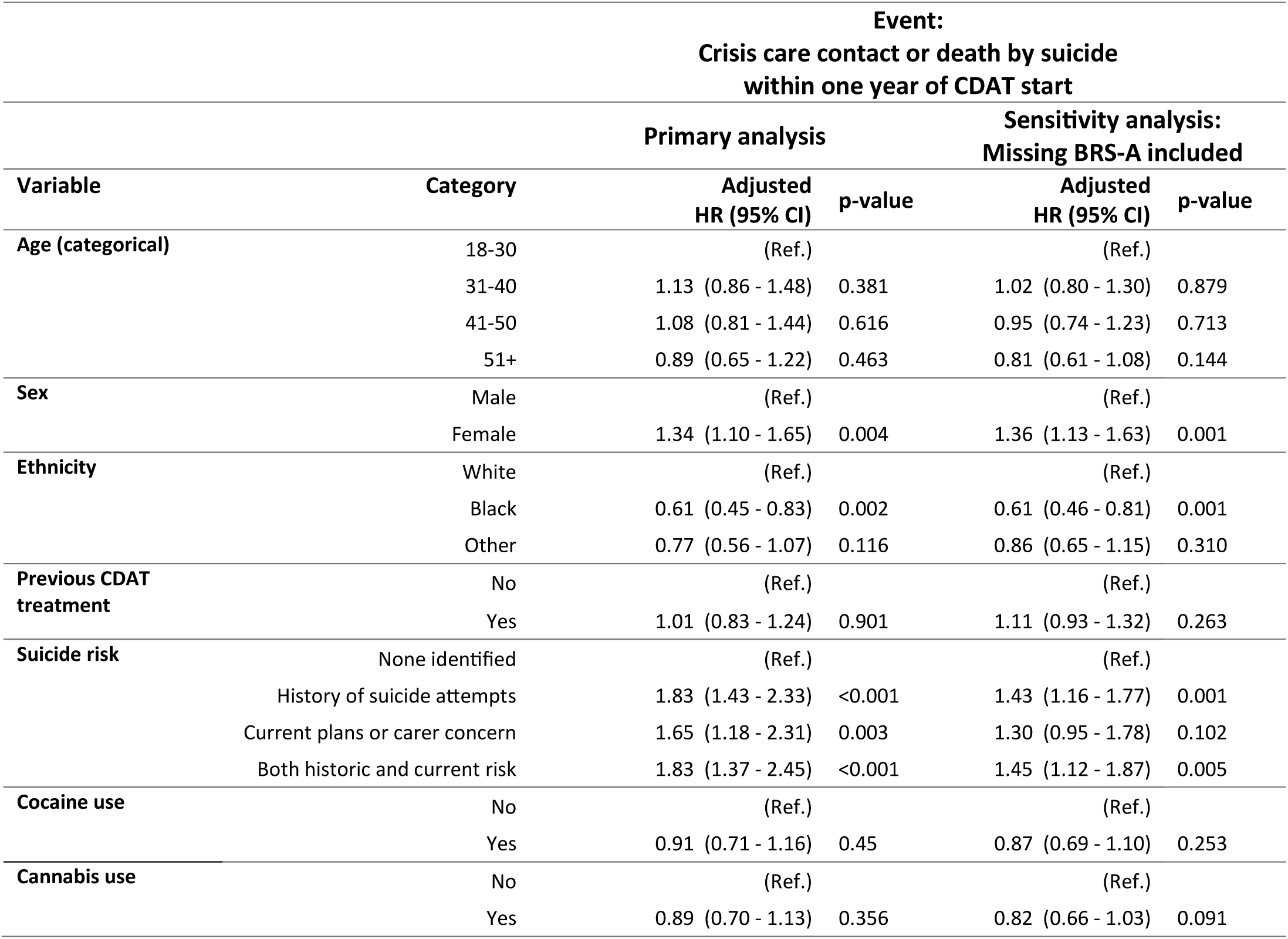
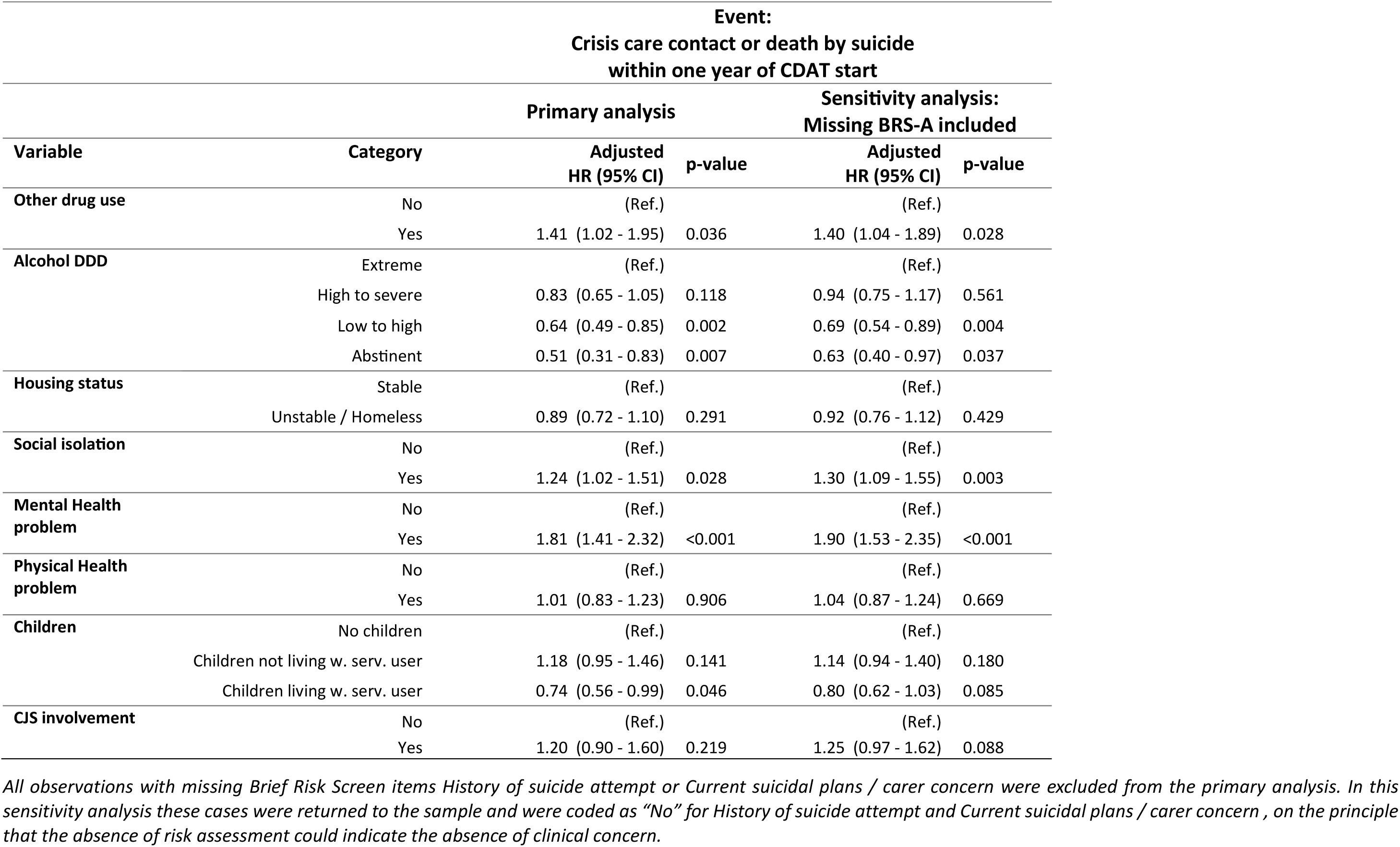
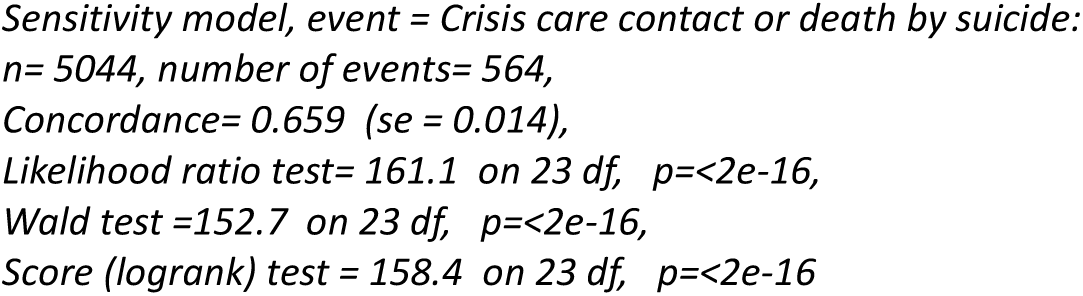
Comparison of primary and sensitivity analyses: Cases with missing BRS-A included.

**Supplementary Table 4.**
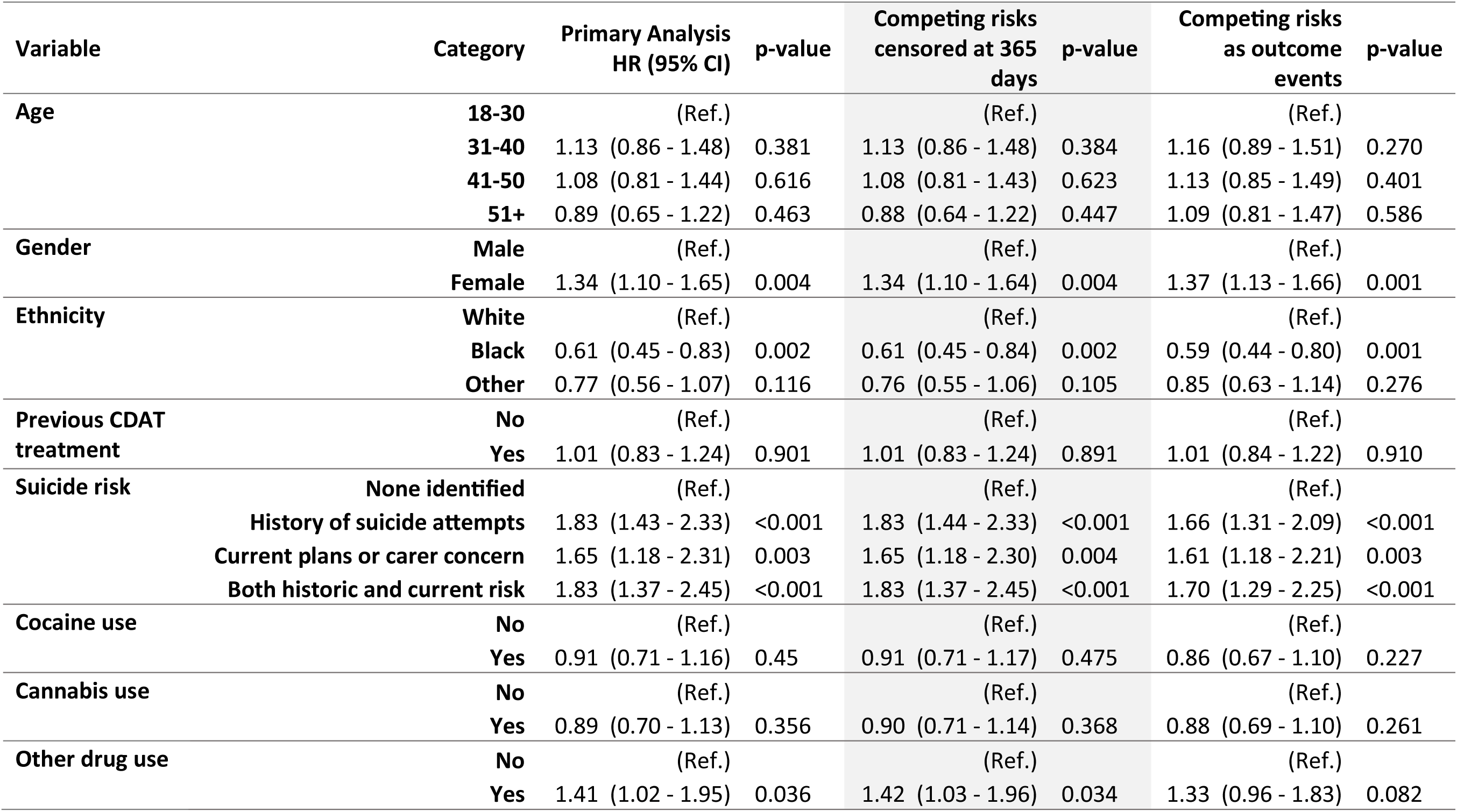
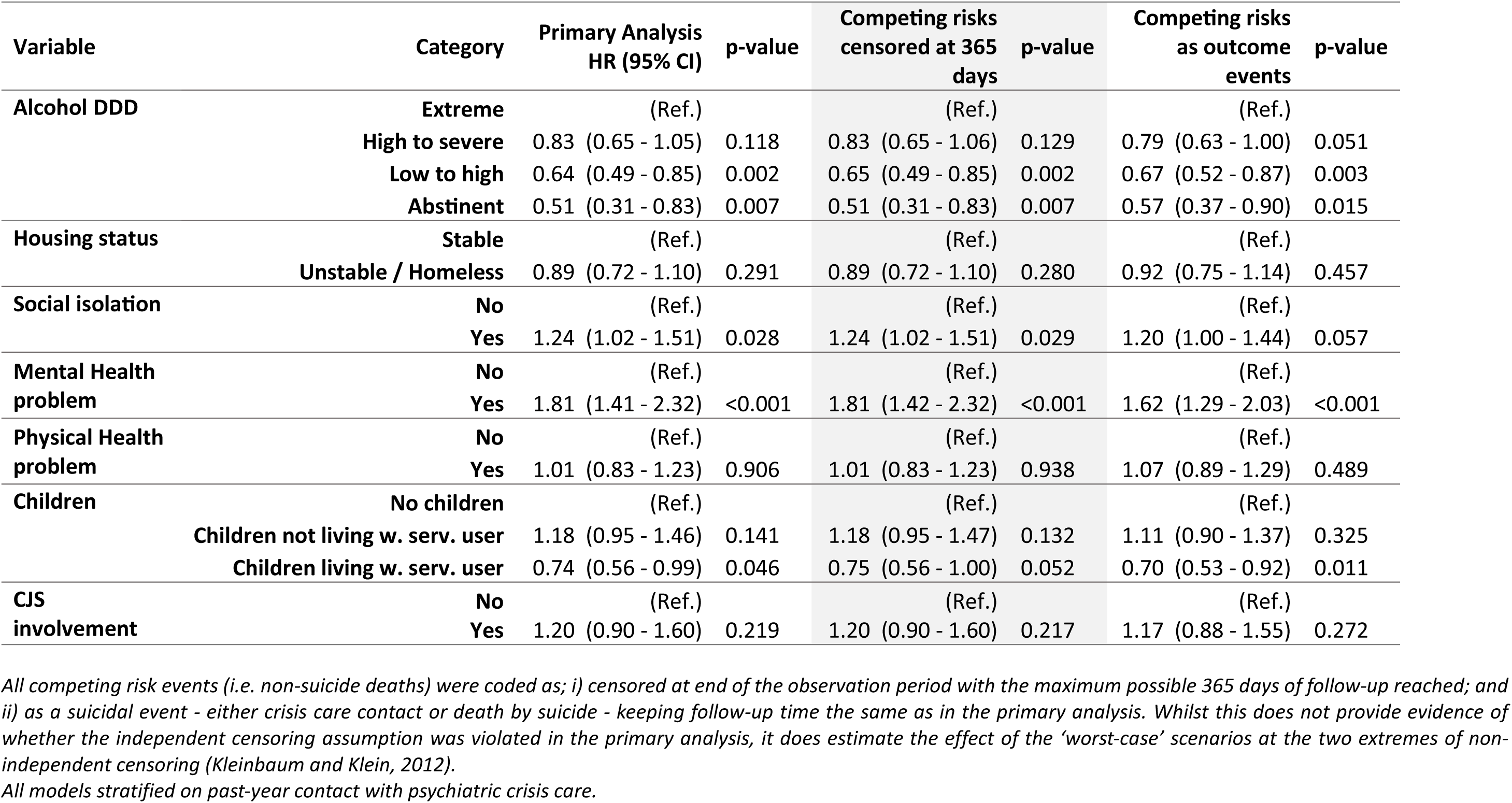
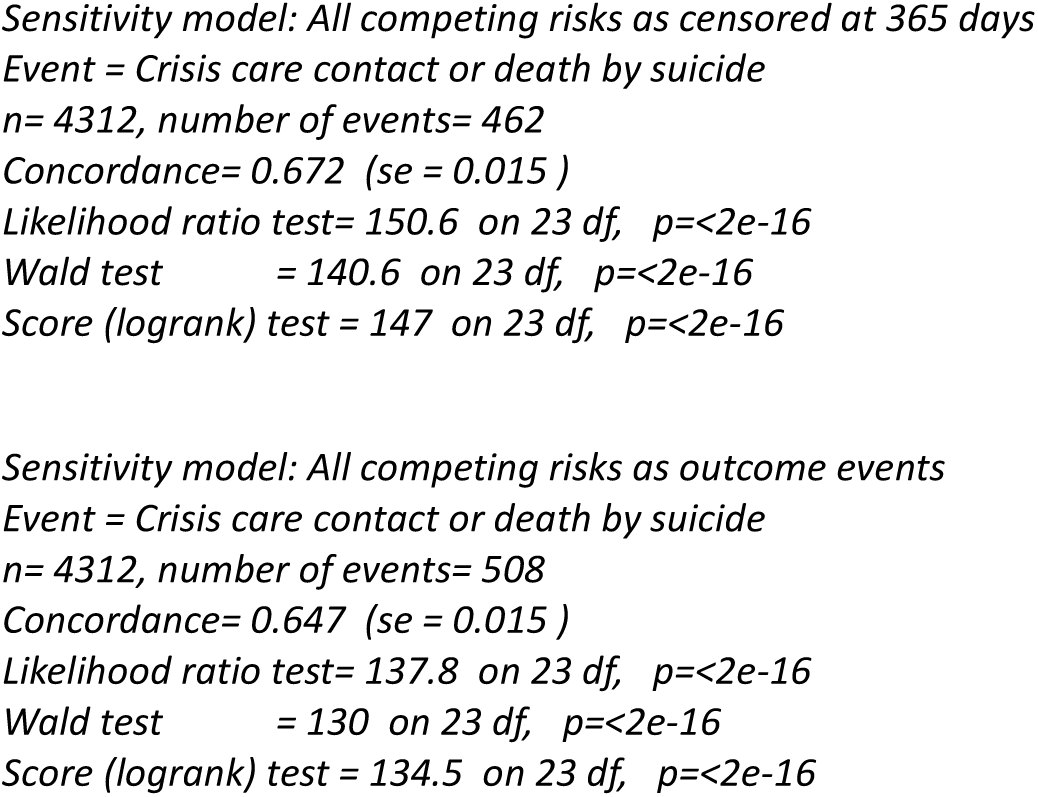
Comparison of primary and sensitivity analyses: Independent censoring assumption.

## References

1. Isaacs JY, Smith MM, Sherry SB, Seno M, Moore ML, Stewart SH. Alcohol use and death by suicide: a meta-analysis of 33 studies. Suicide Life-Threatening Behav [Internet]. 2022 Aug 18;52(4):600–14. Available from: https://onlinelibrary.wiley.com/doi/10.1111/sltb.12846

2. Borges G, Bagge CL, Cherpitel CJ, Conner KR, Orozco R, Rossow I. A meta-analysis of acute use of alcohol and the risk of suicide attempt. Psychol Med. 2017;47(5):949–57.

3. Chesney E, Goodwin GM, Fazel S. Risks of all-cause and suicide mortality in mental disorders: a meta-review. World Psychiatry. 2014;13(2):153–60.

4. Abdul-Rahman AK, Card TR, Grainge MJ, Fleming KM. All-cause and cause-specific mortality rates of patients treated for alcohol use disorders: A meta-analysis. Subst Abus [Internet]. 2018;39(4):509–17. Available from: 10.1080/08897077.2018.1475318

5. Rontziokos H, Deane F. Systematic review of suicidal behaviour in individuals who have attended substance abuse treatment. Int J Ment Health Addict [Internet]. 2019 Dec 28;17(6):1580–98. Available from: http://link.springer.com/10.1007/s11469-018-9994-5

6. Nock MK, Borges G, Bromet EJ, Alonso J, Angermeyer M, Beautrais A, et al. Cross-national prevalence and risk factors for suicidal ideation, plans and attempts. Br J Psychiatry [Internet]. 2008 Feb 2;192(2):98–105. Available from: https://www.cambridge.org/core/product/identifier/S0007125000233886/type/journal_article

7. Darvishi N, Farhadi M, Haghtalab T, Poorolajal J. Alcohol-related risk of suicidal ideation, suicide attempt, and completed suicide: a meta-analysis. Voracek M, editor. PLoS One [Internet]. 2015 May 20;10(5):e0126870. Available from: https://dx.plos.org/10.1371/journal.pone.0126870

8. The National Confidential Inquiry into Suicide and Safety in Mental Health (NCISH). Suicide by people in contact with drug and alcohol services: a national study 2021 to 2022. The University of Manchester; 2024.

9. Hawton K, Lascelles K, Pitman A, Gilbert S, Silverman M. Assessment of suicide risk in mental health practice: shifting from prediction to therapeutic assessment, formulation, and risk management. The Lancet Psychiatry [Internet]. 2022 Nov;9(11):922–8. Available from: 10.1016/S2215-0366(22)00232-2

10. Pisani AR, Murrie DC, Silverman MM. Reformulating suicide risk formulation: from prediction to prevention. Acad Psychiatry [Internet]. 2016 Aug 14;40(4):623–9. Available from: 10.1007/s40596-015-0434-6

11. Webb RT, Qin P, Stevens H, Mortensen PB, Appleby L, Shaw J. National study of suicide in all people with a criminal justice history. Arch Gen Psychiatry [Internet]. 2011 Jun 6;68(6):591. Available from: http://archpsyc.jamanetwork.com/article.aspx?doi=10.1001/archgenpsychiatry.2011.7

12. Ilgen MA, Harris AHS, Moos RH, Tiet QQ. Predictors of a suicide attempt one year after entry into substance use disorder treatment. Alcohol Clin Exp Res [Internet]. 2007 Apr 27;31(4):635–42. Available from: https://onlinelibrary.wiley.com/doi/10.1111/j.1530-0277.2007.00348.x

13. Dehara M, Wells MB, Sjöqvist H, Kosidou K, Dalman C, Sörberg Wallin A. Parenthood is associated with lower suicide risk: a register-based cohort study of 1.5 million Swedes. Acta Psychiatr Scand [Internet]. 2021 Mar 19;143(3):206–15. Available from: https://onlinelibrary.wiley.com/doi/10.1111/acps.13240

14. Haw C, Houston K, Townsend E, Hawton K. Deliberate self-harm patients with alcohol disorders: characteristics, treatment, and outcome. Crisis. 2001;22(3):93–101.

15. Penney A, Mazmanian D, Jamieson J, Black N. Factors associated with recent suicide attempts in clients presenting for addiction treatment. Int J Ment Health Addict [Internet]. 2012 Feb 18;10(1):132–40. Available from: http://link.springer.com/10.1007/s11469-010-9307-0

16. Resko SM, Kruman Mountain S, Browne S, Kondrat DC, Kral M. Suicidal ideation and suicide attempts among women seeking treatment for substance use and trauma symptoms. Health Soc Work [Internet]. 2018 May 1;43(2):76–83. Available from: https://academic.oup.com/hsw/article/43/2/76/4885831

17. Jakubczyk A, Klimkiewicz A, Krasowska A, Kopera M, Sławińska-Ceran A, Brower K, et al. History of sexual abuse and suicide attempts in alcohol-dependent patients. Child Abuse Negl [Internet]. 2014 Sep;38(9):1560–8. Available from: https://linkinghub.elsevier.com/retrieve/pii/S0145213414002233

18. Conner KR, Hesselbrock VM, Meldrum SC, Schuckit MA, Bucholz KK, Gamble SA, et al. Transitions to, and correlates of, suicidal ideation, plans, and unplanned and planned suicide attempts among 3,729 men and women with alcohol dependence. J Stud Alcohol Drugs [Internet]. 2007 Sep 15;68(5):654–62. Available from: https://www.cambridge.org/core/product/identifier/S0022172400000188/type/journal_article

19. Darke S, Williamson A, Ross J, Teesson M. Attempted suicide among heroin users: 12-month outcomes from the Australian Treatment Outcome Study (ATOS). Drug Alcohol Depend [Internet]. 2005 May;78(2):177–86. Available from: https://linkinghub.elsevier.com/retrieve/pii/S0376871604003187

20. Merrall ELC, Bird SM, Hutchinson SJ. Mortality of those who attended drug services in Scotland 1996–2006: record-linkage study. Int J Drug Policy [Internet]. 2012 Jan;23(1):24–32. Available from: 10.1016/j.drugpo.2011.05.010

21. Bogdanowicz KM, Stewart R, Chang CK, Downs J, Khondoker M, Shetty H, et al. Identifying mortality risks in patients with opioid use disorder using brief screening assessment: secondary mental health clinical records analysis. Drug Alcohol Depend [Internet]. 2016;164:82–8. Available from: 10.1016/j.drugalcdep.2016.04.036

22. Wines JD, Saitz R, Horton NJ, Lloyd-Travaglini C, Samet JH. Suicidal behavior, drug use and depressive symptoms after detoxification: a 2-year prospective study. Drug Alcohol Depend [Internet]. 2004 Dec;76S:S21–9. Available from: https://linkinghub.elsevier.com/retrieve/pii/S037687160400211X

23. Preuss UW, Schuckit MA, Smith TL, Danko GP, Bucholz KK, Hesselbrock MN, et al. Predictors and correlates of suicide attempts over 5 years in 1,237 alcohol-dependent men and women. Am J Psychiatry [Internet]. 2003 Jan;160(1):56–63. Available from: http://psychiatryonline.org/doi/abs/10.1176/appi.ajp.160.1.56

24. Darke S, Ross J, Williamson A, Mills KL, Havard A, Teesson M. Patterns and correlates of attempted suicide by heroin users over a 3-year period: findings from the Australian treatment outcome study. Drug Alcohol Depend [Internet]. 2007 Mar;87(2–3):146–52. Available from: https://linkinghub.elsevier.com/retrieve/pii/S0376871606003024

25. Ilgen MA, Jain A, Lucas E, Moos RH. Substance use-disorder treatment and a decline in attempted suicide during and after treatment. J Stud Alcohol Drugs [Internet]. 2007 Jul;68(4):503–9. Available from: http://www.scopus.com/inward/record.url?eid=2-s2.0-34447632189&partnerID=tZOtx3y1

26. Pavarin RM, Sanchini S, Tadonio L, Domenicali M, Caputo F, Pacetti M. Suicide mortality risk in a cohort of individuals treated for alcohol, heroin or cocaine abuse: Results of a follow-up study. Psychiatry Res [Internet]. 2021 Feb;296:113639. Available from: 10.1016/j.psychres.2020.113639

27. Hesse M, Thylstrup B, Seid AK, Skogen JC. Suicide among people treated for drug use disorders: a Danish national record-linkage study. BMC Public Health [Internet]. 2020 Dec 31;20(1):146. Available from: https://bmcpublichealth.biomedcentral.com/articles/10.1186/s12889-020-8261-4

28. Britton PC, Conner KR. Suicide attempts within 12 months of treatment for substance use disorders. Suicide Life-Threatening Behav. 2010;40(1):14–21.

29. OHID. Adult substance misuse treatment statistics 2021 to 2022: report - GOV.UK [Internet]. Office for Health Improvement & Disparities; 2023 [cited 2023 Jan 29]. Available from: https://www.gov.uk/government/statistics/substance-misuse-treatment-for-adults-statistics-2021-to-2022/adult-substance-misuse-treatment-statistics-2021-to-2022-report

30. Perera G, Broadbent M, Callard F, Chang C-K, Downs J, Dutta R, et al. Cohort profile of the South London and Maudsley NHS Foundation Trust Biomedical Research Centre (SLaM BRC) Case Register: current status and recent enhancement of an Electronic Mental Health Record-derived data resource. BMJ Open [Internet]. 2016 Mar 1;6(3):e008721. Available from: https://bmjopen.bmj.com/lookup/doi/10.1136/bmjopen-2015-008721

31. Marsden J, Darke S, Hall W, Hickman M, Holmes J, Humphreys K, et al. Mitigating and learning from the impact of COVID-19 infection on addictive disorders. Addiction [Internet]. 2020 Jun 28;115(6):1007–10. Available from: https://onlinelibrary.wiley.com/doi/10.1111/add.15080

32. Mathers BM, Degenhardt L, Bucello C, Lemon J, Wiessing L, Hickman M. Mortality among people who inject drugs: a systematic review and meta-analysis. Bull World Health Organ [Internet]. 2013 Feb 1;91(2):102–23. Available from: https://www.ncbi.nlm.nih.gov/pmc/articles/PMC3605003/pdf/BLT.12.108282.pdf/

33. Lewer D, Brothers TD, Van Hest N, Hickman M, Holland A, Padmanathan P, et al. Causes of death among people who used illicit opioids in England, 2001–18: a matched cohort study. Lancet Public Heal [Internet]. 2022 Feb;7(2):e126–35. Available from: 10.1016/S2468-2667(21)00254-1

34. Bohnert ASB, Roeder K, Ilgen MA. Unintentional overdose and suicide among substance users: a review of overlap and risk factors. Drug Alcohol Depend [Internet]. 2010;110(3):183–92. Available from: 10.1016/j.drugalcdep.2010.03.010

35. PHE. National Drug Treatment Monitoring System (NDTMS): Reference data: Core dataset P (V15.01) [Internet]. London, UK: Public Health England; 2019. Available from: https://assets.publishing.service.gov.uk/media/5e662020d3bf7f1091979e91/NDTMS_Reference_Data_CDS-P_V15.01_docx.pdf

36. Marsden J, Farrell M, Bradbury C, Dale-Perera A, Eastwood B, Roxburgh M, et al. Development of the treatment outcomes profile. Addiction [Internet]. 2008 Sep;103(9):1450–60. Available from: https://onlinelibrary.wiley.com/doi/10.1111/j.1360-0443.2008.02284.x

37. Peacock A, Eastwood B, Jones A, Millar T, Horgan P, Knight J, et al. Effectiveness of community psychosocial and pharmacological treatments for alcohol use disorder: a national observational cohort study in England. Drug Alcohol Depend [Internet]. 2018 May;186(June 2017):60–7. Available from: 10.1016/j.drugalcdep.2018.01.019

38. NICE. Alcohol-use disorders: diagnosis, assessment and management of harmful drinking (high-risk drinking) and alcohol dependence (CG115) [Internet]. London, UK: The British Psychological Society & The Royal College of Psychiatrists; 2011. Available from: https://www.nice.org.uk/guidance/cg115/evidence/full-guideline-136423405

39. NCISH. The National Confidential Inquiry into Suicide and Safety in Mental Health: Safer services : a toolkit for specialist mental health services and primary care [Internet]. Manchester: University of Manchester; 2022. Available from: https://documents.manchester.ac.uk/display.aspx?DocID=40697

40. R Core Team. R: A Language and Environment for Statistical Computing. Vienna: R Foundation for Statistical Computing; 2023.

41. Kleinbaum DG, Klein M. Survival analysis. 3rd Ed. Statistics for Biology and Health. Springer; 2012.

42. Yoshimasu K, Kiyohara C, Miyashita K. Suicidal risk factors and completed suicide: meta-analyses based on psychological autopsy studies. Environ Health Prev Med [Internet]. 2008 Sep 19;13(5):243–56. Available from: http://link.springer.com/10.1007/s12199-008-0037-x

43. Robins JE, Morley KI, Hayes RD, Ross KR, Pritchard M, Curtis V, et al. Alcohol dependence and heavy episodic drinking are associated with different levels of risk of death or repeat emergency service attendance after a suicide attempt. Drug Alcohol Depend [Internet]. 2021 Jul;224:108725. Available from: 10.1016/j.drugalcdep.2021.108725

44. Robins JE, Kalk NJ, Ross KR, Pritchard M, Curtis V, Morley KI. The association of acute alcohol use and dynamic suicide risk with variation in onward care after psychiatric crisis. Drug Alcohol Rev [Internet]. 2021 Mar 10;40(3):499–508. Available from: https://onlinelibrary.wiley.com/doi/10.1111/dar.13231

45. Hunt IM, Richards N, Bhui K, Ibrahim S, Turnbull P, Halvorsrud K, et al. Suicide rates by ethnic group among patients in contact with mental health services: an observational cohort study in England and Wales. The Lancet Psychiatry [Internet]. 2021 Dec;8(12):1083–93. Available from: 10.1016/S2215-0366(21)00354-0

46. Canfield M, Norton S, Downs J, Gilchrist G. Parental status and characteristics of women in substance use treatment services: analysis of electronic patient records. J Subst Abuse Treat [Internet]. 2021;127(108365). Available from: 10.1016/j.jsat.2021.108365

47. Parmar M, Ma R, Attygalle S, Mueller C, Stubbs B, Stewart R, et al. Associations between loneliness and acute hospitalisation outcomes among patients receiving mental healthcare in South London: a retrospective cohort study. Soc Psychiatry Psychiatr Epidemiol [Internet]. 2022 Feb 20;57(2):397–410. Available from: 10.1007/s00127-021-02079-9

48. Drinkwater C, Wildman J, Moffatt S. Social prescribing. BMJ [Internet]. 2019 Mar 28;364:l1285. Available from: https://www.bmj.com/lookup/doi/10.1136/bmj.l1285

49. Ross J, Darke S, Kelly E, Hetherington K. Suicide risk assessment practices: a national survey of generalist drug and alcohol residential rehabilitation services. Drug Alcohol Rev [Internet]. 2012 Sep 27;31(6):790–6. Available from: https://onlinelibrary.wiley.com/doi/10.1111/j.1465-3362.2012.00437.x

50. Zisman S, O’Brien A. A retrospective cohort study describing six months of admissions under Section 136 of the Mental Health Act; the problem of alcohol misuse. Med Sci Law [Internet]. 2015 Jul 24 [cited 2018 Feb 12];55(3):216–22. Available from: http://journals.sagepub.com/doi/10.1177/0025802414538247

51. Yates K, Lång U, Cederlöf M, Boland F, Taylor P, Cannon M, et al. Association of Psychotic Experiences With Subsequent Risk of Suicidal Ideation, Suicide Attempts, and Suicide Deaths. JAMA Psychiatry [Internet]. 2019 Feb 1;76(2):180. Available from: http://archpsyc.jamanetwork.com/article.aspx?doi=10.1001/jamapsychiatry.2018.3514

52. Appleby L, Turnbull P, Kapur N, Gunnell D, Hawton K. New standard of proof for suicide at inquests in England and Wales. BMJ [Internet]. 2019;366. Available from: doi:10.1136/bmj.l4745

